# Association of plasma proteins with rate of cognitive decline and dementia: 20-year follow-up of the Whitehall II and ARIC cohort studies

**DOI:** 10.1101/2020.11.18.20234070

**Authors:** Joni V. Lindbohm, Nina Mars, Keenan A. Walker, Archana Singh-Manoux, Gill Livingston, Eric J. Brunner, Pyry N. Sipilä, Kalle Saksela, Jane E. Ferrie, Ruth Lovering, Stephen A. Williams, Aroon D. Hingorani, Rebecca F. Gottesman, Henrik Zetterberg, Mika Kivimäki

**Affiliations:** Department of Epidemiology and Public Health, University College London, 1-19 Torrington Place, London, UK; Clinicum, Department of Public Health, University of Helsinki, P.O. Box 41, FI-00014 Helsinki, Finland; Institute for Molecular Medicine Finland (FIMM), HiLIFE, University of Helsinki, Helsinki, Finland; Department of Neurology, The Johns Hopkins University, Baltimore, MD, USA; Université de Paris, Inserm U1153, Epidemiology of Ageing and Neurodegenerative diseases, Paris, France; Division of Psychiatry, University College London, London, UK; Camden and Islington Foundation Trust, London, UK; Department of Virology, University of Helsinki and HUSLAB, Helsinki University Hospital, Helsinki, Finland; Bristol Medical School (PHS), University of Bristol, Bristol, UK; Functional Gene Annotation, Institute of Cardiovascular Science, University College London, London, UK; SomaLogic, Inc. Boulder, CO, USA; Institute of Cardiovascular Science, University College London, London, UK; University College London, British Heart Foundation Research Accelerator, London, UK; Health Data Research UK, London, UK; Department of Neurodegenerative Disease and UK Dementia Research Institute, University College London, UK; Department of Psychiatry and Neurochemistry, Institute of Neuroscience and Physiology, The Sahlgrenska Academy, University of Gothenburg, Sweden; Clinical Neurochemistry Laboratory, Sahlgrenska University Hospital, Mölndal, Sweden

## Abstract

The role of circulating proteins in Alzheimer’s disease and related dementias is unknown. Using a follow-up of two decades, 4953 plasma proteins, and discovery (Whitehall II) and replication cohort (ARIC), we examined plasma proteins associated with cognitive decline rate and dementia. After replication and adjustment for known dementia risk factors, fifteen proteins were associated with cognitive decline rate and dementia. None of these were amyloid, tau, or neurofilament-related proteins. Currently approved medications can target five of the proteins. The results support systemic pathogenesis of dementias, may aid in early diagnosis, and suggest potential targets for drug development.

## Introduction

Alzheimer’s disease and related dementias pose an increasing challenge with considerable costs to the individual and to health and social care services.^1^ Until recently, amyloid beta and tau proteins have dominated pathophysiological models of dementia aetiology.^2^ However, numerous prevention and treatment trials targeting these biomarkers have failed.^3, 4^ In addition, longitudinal studies suggest that most cognitively normal amyloid-positive people never develop clinical dementia.^5^ Given this and the high prevalence of mixed types of dementia in the general population, there is a need to expand research on early biomarkers for dementia beyond amyloid and tau.

Causes of dementia are increasingly thought to be systemic.^6, 7^ Recent development of scalable platforms allows simultaneous assessment thousands of circulating proteins.^8, 9^ These have the potential to identify drivers of dementia, and move the field beyond amyloid and tau. For several reasons, circulating proteins are promising targets for biomarker and drug discovery. Proteins are regulators and effectors of biological processes and also imprints of the effects of genes, the environment, age, current co-morbidities, behaviours, and medications, all of which may affect dementia development.^10^ Proteins are also highly druggable as they can be targeted by monoclonal antibodies, small molecule drugs, or proteolysis-targeting chimaeras.^11-14^ Indeed, of all currently approved medications, approximately 96% target proteins.^11^ Animal studies also support a causal role of circulating plasma proteins in neurodegeneration. Plasma from young mice, via injection or through parabiosis (joining two animals so they share blood circulation) has been shown to restore memory and stimulate synaptic plasticity in the aged mouse hippocampus.^15-17^ In humans, multiple circulating proteins have been linked with dementia, but the studies to date have been mainly cross-sectional, based on small samples, or lacked replication.^18^

In this report from two large cohort studies, the British Whitehall II and the US Atherosclerosis Risk in Communities (ARIC) study, we used SomaScan technology to examine 4953 plasma proteins, including 22 related to amyloid, tau, or neurofilaments, as risk factors for cognitive decline and dementia. To determine what role proteins play in the early stages of neurodegeneration when dementia may still be preventable, we assessed plasma proteins in late middle age and followed subsequent cognitive decline and dementia over two decades. Thus, our study design explicitly takes into consideration the long preclinical phase of dementia.

## Results

In the random sample of 2274 participants in the Whitehall II discovery cohort, mean age was 56.1 (SD 5.9) and 1653 (73.0%) were men (**Table 1 and Figure 1**). During a mean follow-up of 20.4 years, 106 individuals developed dementia (36 were Alzheimer’s, 19 vascular, and 51 mixed or other dementias). **Figure 2, part A** shows that those who developed dementia had steeper cognitive decline slopes than participants who remained dementia-free. After FDR correction, 246 of the 4953 proteins were associated with increased rate of cognitive decline (**Figure 2, part B**). None of the amyloid-, tau-, or neurofilament-related proteins were associated with accelerated cognitive decline after FDR correction (**Supplementary Table 1 and Supplementary Table 2)**.

**Table 1.**
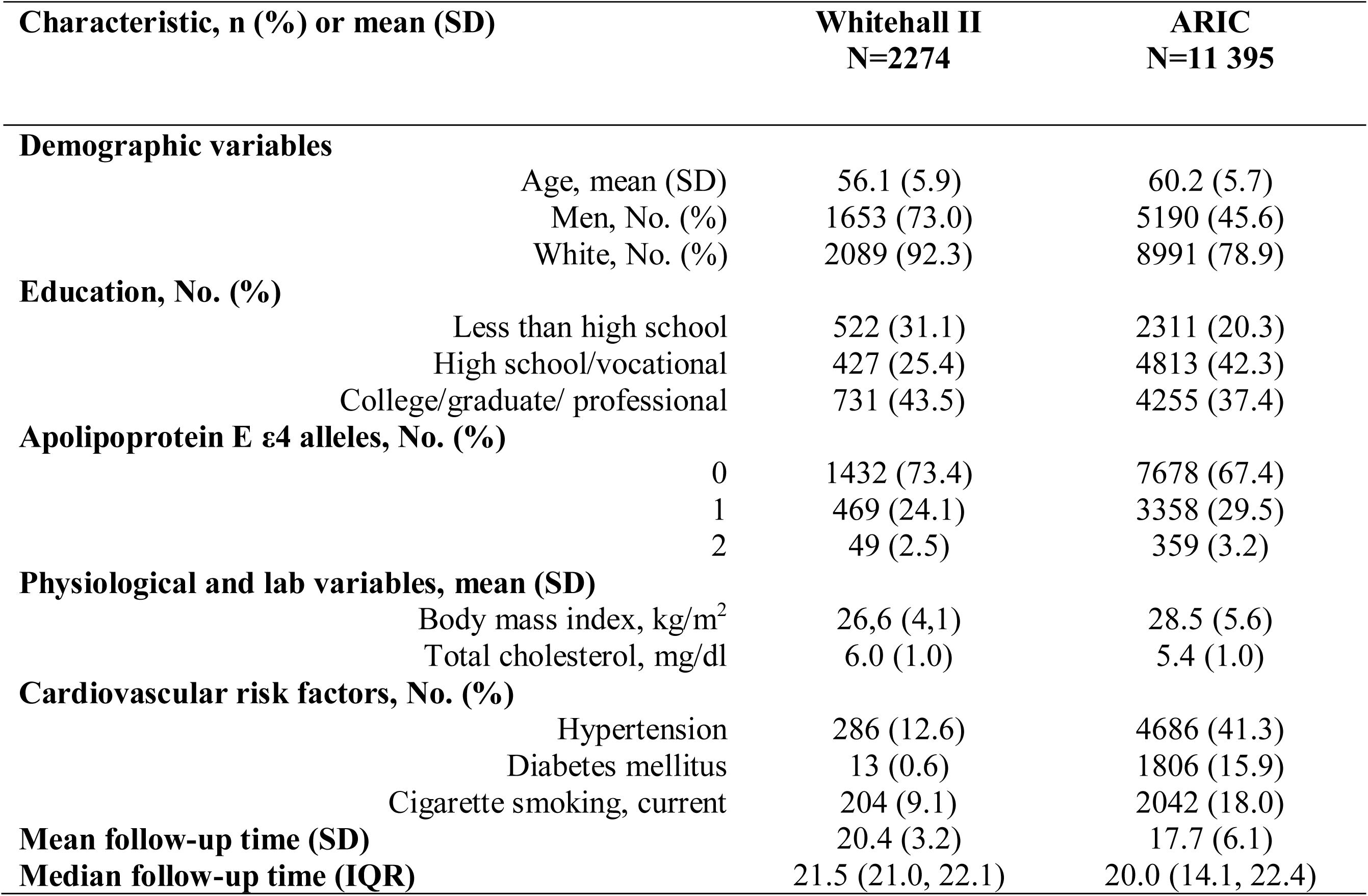
Whitehall and ARIC participant characteristics at the time of blood collection for protein measurement. Values are displayed as means (SD) for continuous variables, frequency (column percentages) for categorical variables, and interquartile range (IQR) for follow-up time.

| Characteristic, n (%) or mean (SD) |  | Whitehall II<br>N=2274 | ARIC<br>N=11 395 |
| --- | --- | --- | --- |
| **Demographic variables** |  |  |  |
|  | Age, mean (SD) | 56.1 (5.9) | 60.2 (5.7) |
|  | Men, No. (%) | 1653 (73.0) | 5190 (45.6) |
|  | White, No. (%) | 2089 (92.3) | 8991 (78.9) |
| **Education, No. (%)** |  |  |  |
|  | Less than high school | 522 (31.1) | 2311 (20.3) |
|  | High school/vocational | 427 (25.4) | 4813 (42.3) |
|  | College/graduate/ professional | 731 (43.5) | 4255 (37.4) |
| **Apolipoprotein E ε4 alleles, No. (%)** |  |  |  |
|  | 0 | 1432 (73.4) | 7678 (67.4) |
|  | 1 | 469 (24.1) | 3358 (29.5) |
|  | 2 | 49 (2.5) | 359 (3.2) |
| **Physiological and lab variables, mean (SD)** |  |  |  |
|  | Body mass index, kg/m<sup>2</sup> | 26.6 (4.1) | 28.5 (5.6) |
|  | Total cholesterol, mg/dl | 6.0 (1.0) | 5.4 (1.0) |
| **Cardiovascular risk factors, No. (%)** |  |  |  |
|  | Hypertension | 286 (12.6) | 4686 (41.3) |
|  | Diabetes mellitus | 13 (0.6) | 1806 (15.9) |
|  | Cigarette smoking, current | 204 (9.1) | 2042 (18.0) |
| **Mean follow-up time (SD)** |  | 20.4 (3.2) | 17.7 (6.1) |
| **Median follow-up time (IQR)** |  | 21.5 (21.0, 22.1) | 20.0 (14.1, 22.4) |

**Table 2.** Associations between the proteins and dementia related pathologies, existing evidence of these associations, and current medications that can influence protein function or level

| Protein | Immune system | BBB* | Vascular pathology | Central insulin resistance | Previous studies | Medications (indication) |
| --- | --- | --- | --- | --- | --- | --- |
| N-terminal pro-BNP | - | - | 1 | - | Prospective cohort<sup>27,28,76,77</sup> | Antihypertensive medications (hypertension)<sup>74</sup> |
| CDCP1 | 1 | 1 | - | - | None | Itolizumab<sup>88</sup> (psoriasis) |
| MIC-1 | 1 | 1 | 1 | - | Case control<sup>22,23</sup> | - |
| CRDL1 | - | - | 1 | - | None | - |
| RNAS6 | 1 | - | - | - | None | - |
| SAP3 | - | 1 | 1 | - | Case-control<sup>19</sup> | - |
| HE4 | 1 | 1 | 1 | - | None | - |
| TIMP-4 | 1 | 1 | 1 | - | Case-control<sup>23</sup> | - |
| IGFBP-7 | - | - | 1 | 1 | Case-control<sup>20</sup> | Intranasal insulin, metformin, and GLP-1 receptor agonists<sup>86</sup> (Alzheimer's disease, diabetes) |
| OPG | 1 | 1 | 1 | - | Case-control<sup>25</sup> | Atorvastatin, metformin, glitazones<sup>56,89,90</sup> (atherosclerotic cardiovascular diseases, diabetes) |
| SIGLEC-7 | 1 | - | - | - | None | - |
| SVEP1 | 1 | 1 | 1 | - | None | - |
| TREM2 | 1 | - | - | - | Prospective cohort<sup>27</sup> | - |
| NPS-PLA2 | 1 | - | 1 | - | Case-control<sup>27</sup> | Varespladib and Varespladib Methyl<sup>87</sup> (cardiovascular and inflammatory diseases) |
| MARCKSL1 | 1 | 1 | - | - | None | - |
| Total | 11 | 8 | 10 | 1 |  |  |

\*BBB = blood brain barrier

**Figure 1.**
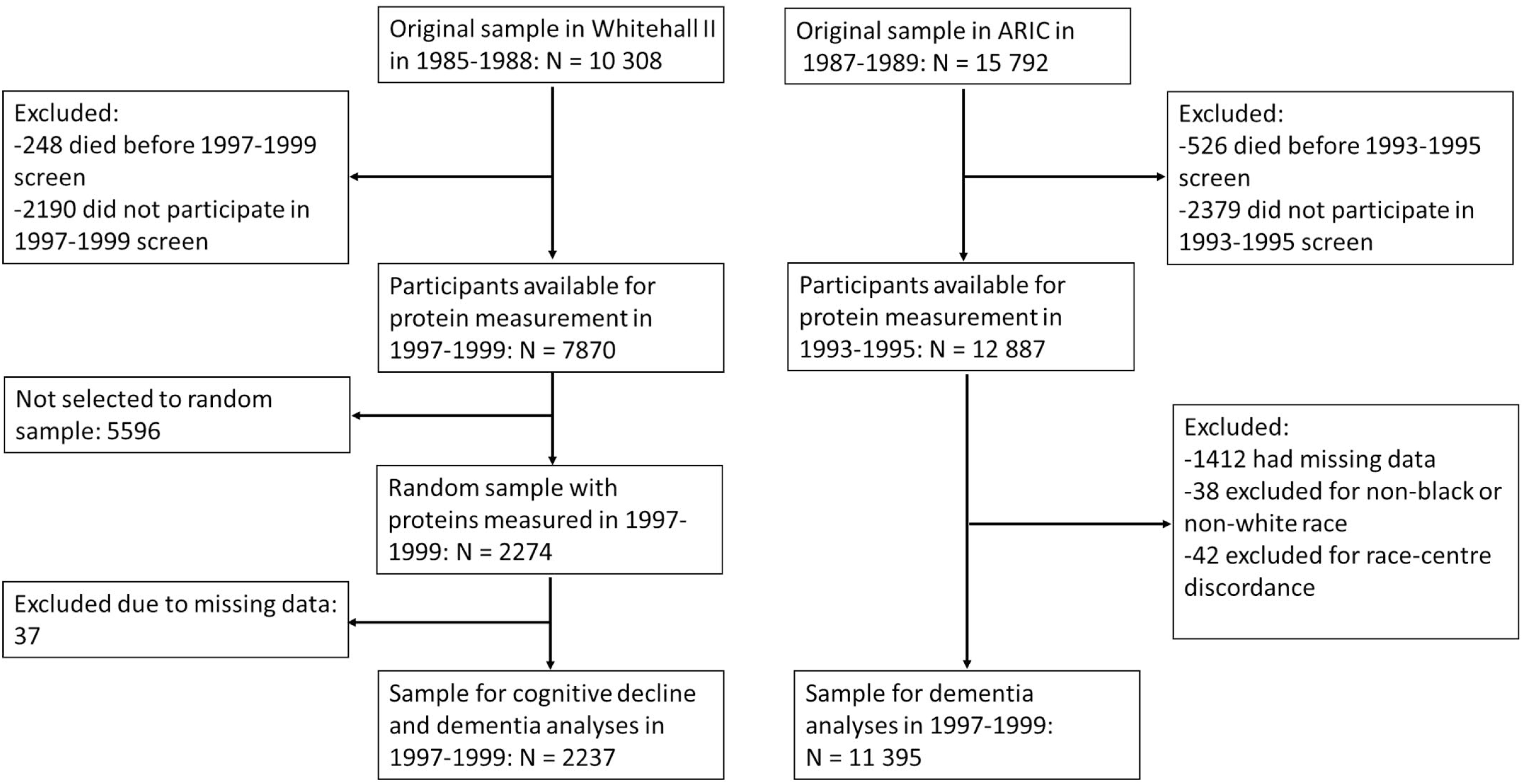
Flowchart of the study design.

**Figure 2.**
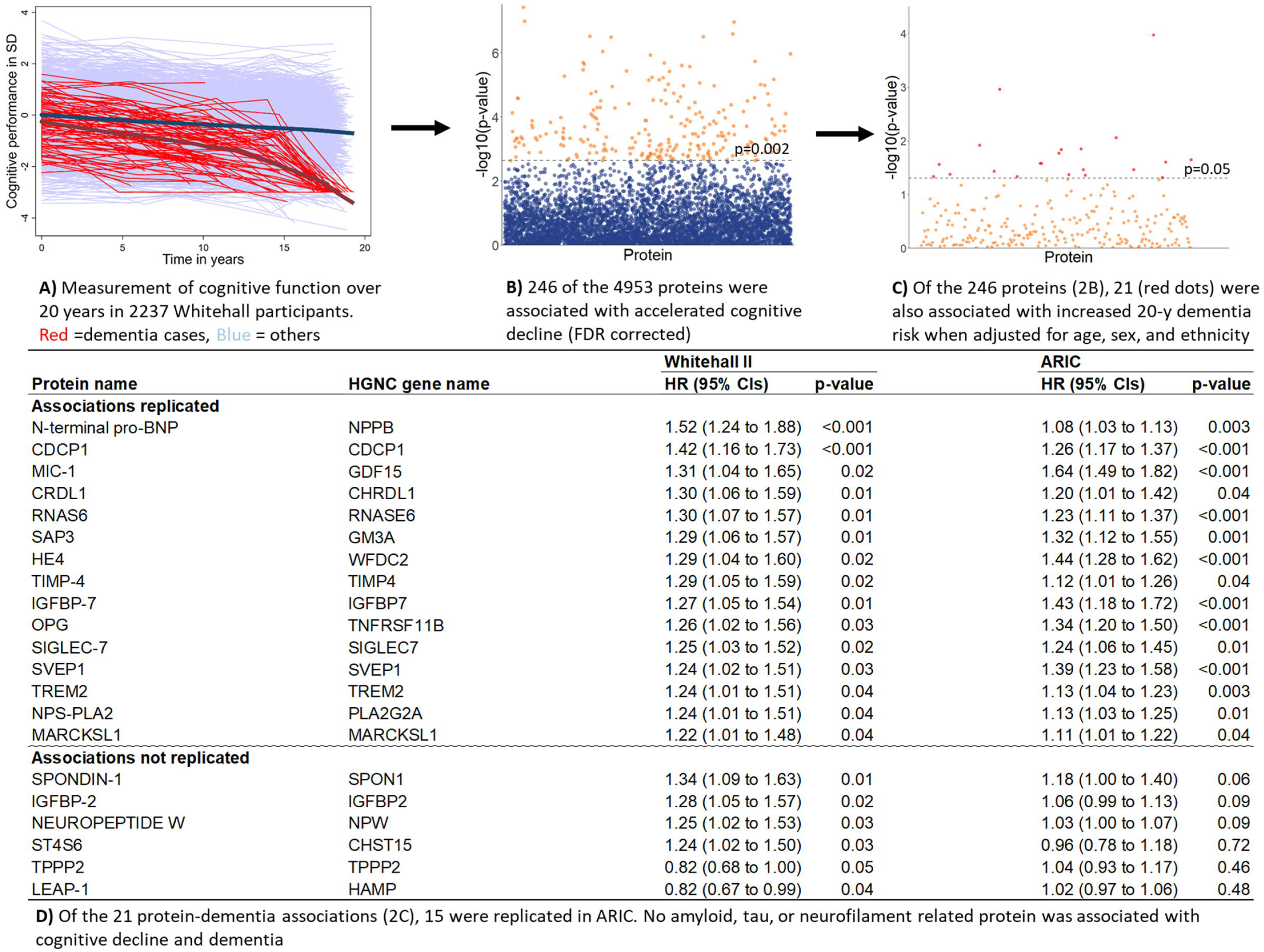
Summary of the three steps in protein analysis.

Of the 246 proteins, 21 were also associated with incident dementia in the Whitehall II study (**Figure 2, part C)**. Thus, replication analyses in ARIC were based on 21 proteins. In this cohort, 1942 of the 11 395 participants developed dementia during a mean follow-up of 17.4 years. Fifteen of the 21 proteins associated with cognitive decline and dementia in the Whitehall II study were also associated with dementia in the ARIC cohort (**Figure 2, part D**) with the hazard ratios ranging between 1.08 and 1.64 in age, sex, and ethnicity/race adjusted analyses.

In additional analyses based on the Whitehall data, the 15 protein-dementia associations changed little after further adjustment for age, sex, ethnicity, SBP, antihypertensive medication, BMI, alcohol consumption, smoking, diabetes, socio-economic, and *APOE* status (**Supplementary Figures 1 and 2**) and taking into account competing risk of death (**Supplementary Figure 3**). A cognitive domain specific analysis suggested that phonemic fluency and executive functioning mostly drove the associations with increased rate of cognitive decline (**Supplementary Tables 3-7**). There were no strong correlations between the 15 proteins (all correlations ≤|0.53|, **Supplementary Figure 4**).

The 15 proteins were mostly expressed in tissues other than the brain (**Figure 3 and Supplementary Table 8**). Of the proteins, 14 were secreted and one was a cell membrane protein. All of them were related to one or more biological systems that are relevant to dementia aetiology: dysfunction of the immune system and blood-brain barrier (BBB), vascular pathology, and central insulin resistance (**Table 1**). For five proteins, a medication that can influence the protein’s function (NPS-PLA2, CDCP1, and IGFBP-7) or reduce its levels (N-terminal pro-BNP, and OPG) is available. All these medications have passed phase III trials and are used to treat diabetes, inflammatory- or cardiovascular diseases (**Supplementary Table 9**).

**Figure 3.**
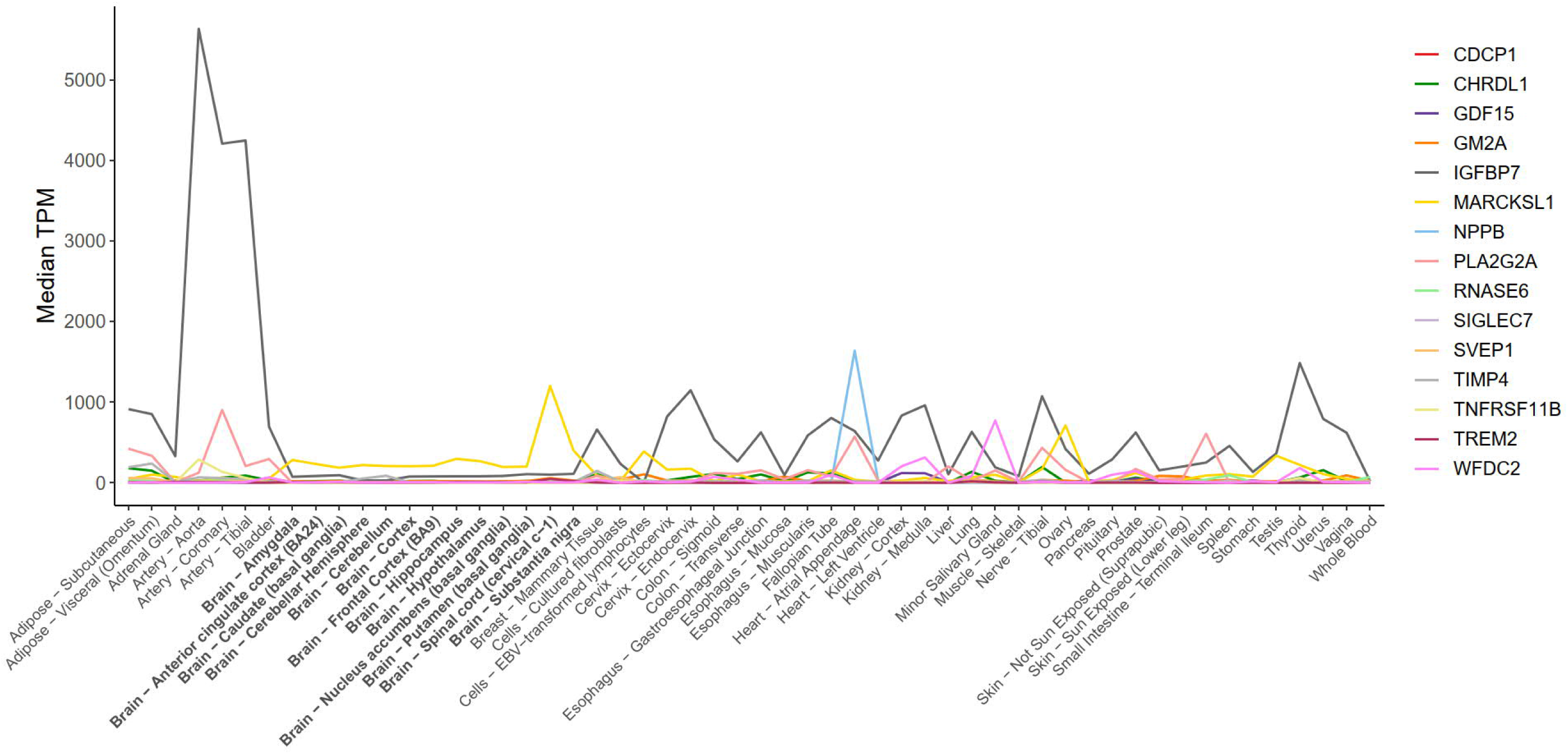
Expression profile in transcripts per million (TPM) of the genes coding the 15 proteins associated with cognitive decline and dementia that survived multivariate adjustment, competing risks, and replication.

## Discussion

This study identified 15 plasma proteins that were associated with increased rate of cognitive decline and increased 20-year risk of dementia in the British Whitehall II study. Of these 14 were secreted (N-terminal pro-BNP, CDCP1, MIC-1, CRDL1, RNAS6, SAP3, HE4, TIMP-4, IGFBP-7, OPG, SVEP1, TREM2, NPS-PLA2, MARCKSL1), and one was a cell membrane protein (SIGLEC-7), all mostly expressed outside central nervous system. The associations with dementia were replicated in an independent US cohort study, ARIC; were robust to adjustments for known dementia risk factors, such as hypertension, smoking, diabetes, and *APOE* status; and were not explained by competing risk of death. Of the 15 proteins, 5 (CDCP1, NPS-PLA2, IGFBP-7, N-terminal pro-BNP, and OPG) are modifiable using existing medications indicated for conditions other than dementias. To our knowledge, this is the largest proteome-wide study with replicated results on the long-term associations between plasma proteins, cognitive decline and risk of dementia.

Our database search on protein expression profiles found that genes encoding the 15 proteins are mainly expressed in tissues other the brain. This suggests that the elevated circulating protein levels in individuals at increased dementia risk are unlikely to represent markers of neurodegeneration resulting from proteins leaking from damaged cells in the central nervous system. In contrast, the expression profiles are consistent with the hypothesis that the identified proteins relate to systemic processes that increase dementia risk.

Of the 15 protein-dementia associations, seven (SVEP1, HE4, CDCP1, SIGLEC-7, MARCKSL1, CRDL1, and RNAS6) are novel, six (SAP3, NPS-PLA2, IGFBP-7, MIC-1, TIMP-4, OPG) have been previously reported in case-control studies^19-26^ and two (TREM2, N-terminal pro-BNP) in prospective cohort studies,^27-29^ all showing the same direction as in this study.

Eleven of the proteins (six novel and five previously identified) are associated with immune response (**Figures 4A and B**). Of these, NPS-PLA2, TREM2, MIC-1, OPG and TIMP-4,^21, 24, 25, 27, 30-35^ have been considered to increase dementia risk due to involvement in activation, adhesion, survival promotion, and infiltration of neutrophils and macrophages. We identified two additional proteins, SVEP1 and MARCKSL1, which may also act in these processes.^36, 37^ SVEP1 can also activate the complement system, which, if prolonged, can be detrimental to endothelial cells.^38^ Leakage of complement through damaged endothelium into the CNS, in turn, can cause the synapse loss in the hippocampus observed in dementia.^39^ This is supported by a recent consortium-based case-control study that showed an association between complement related sCR1, factor B, and factor H and Alzheimer′s disease.^40^ The elevated levels of SVEP1 among asymptomatic individuals who developed dementia in the present study suggests that SVEP1 might play a role in abnormal complement activation that predisposes to dementia.^41^

**Figure 4.**
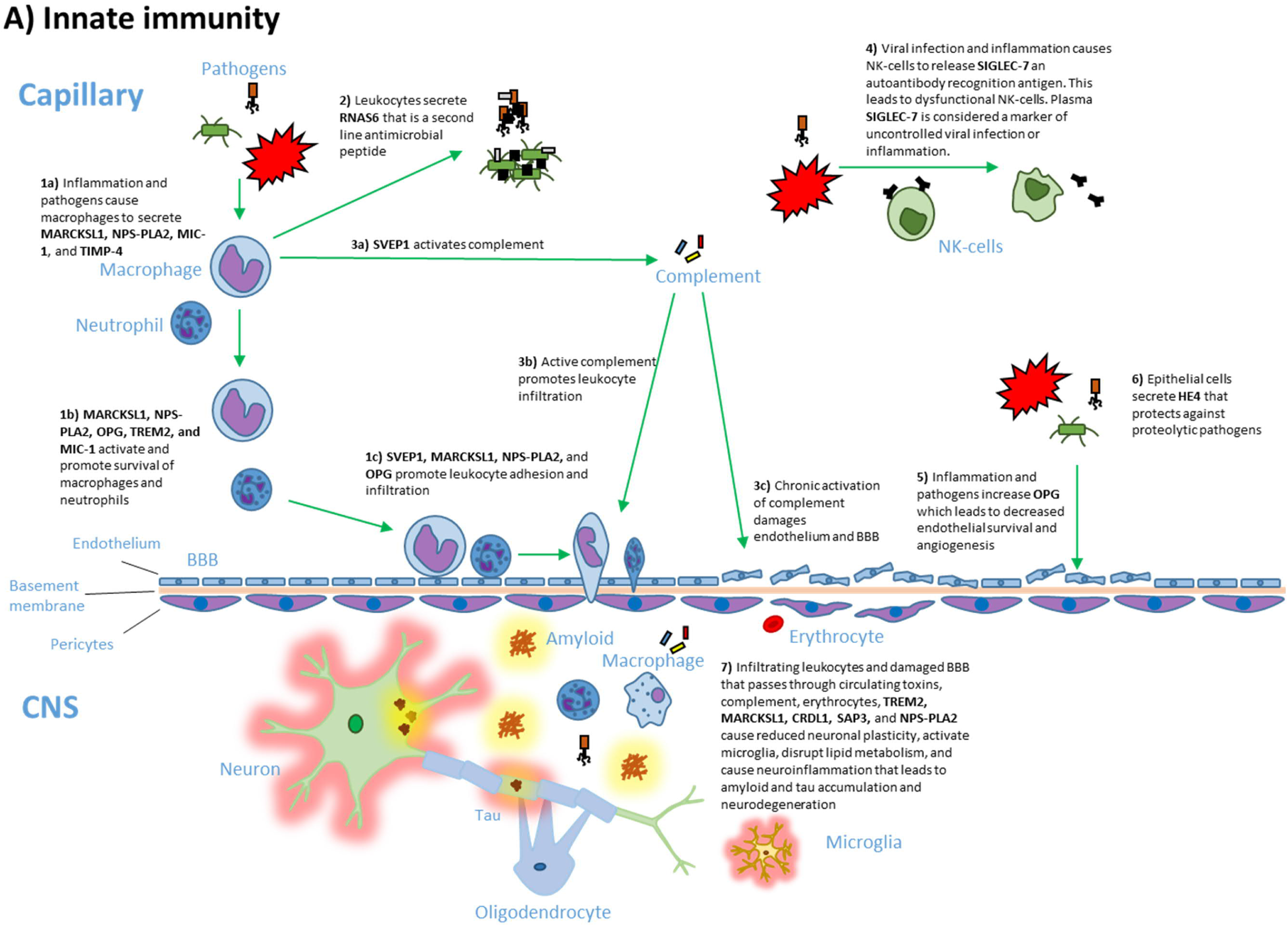
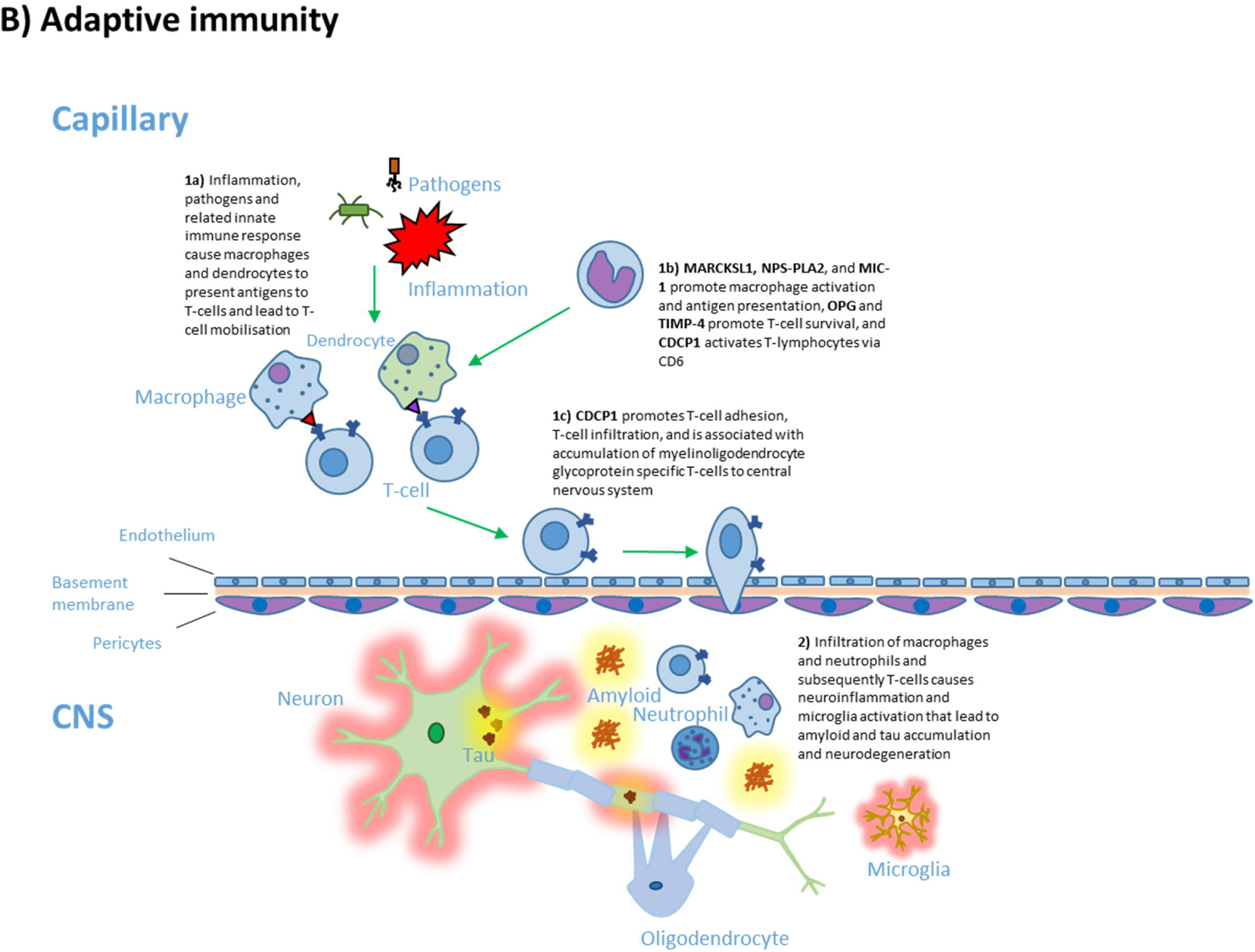
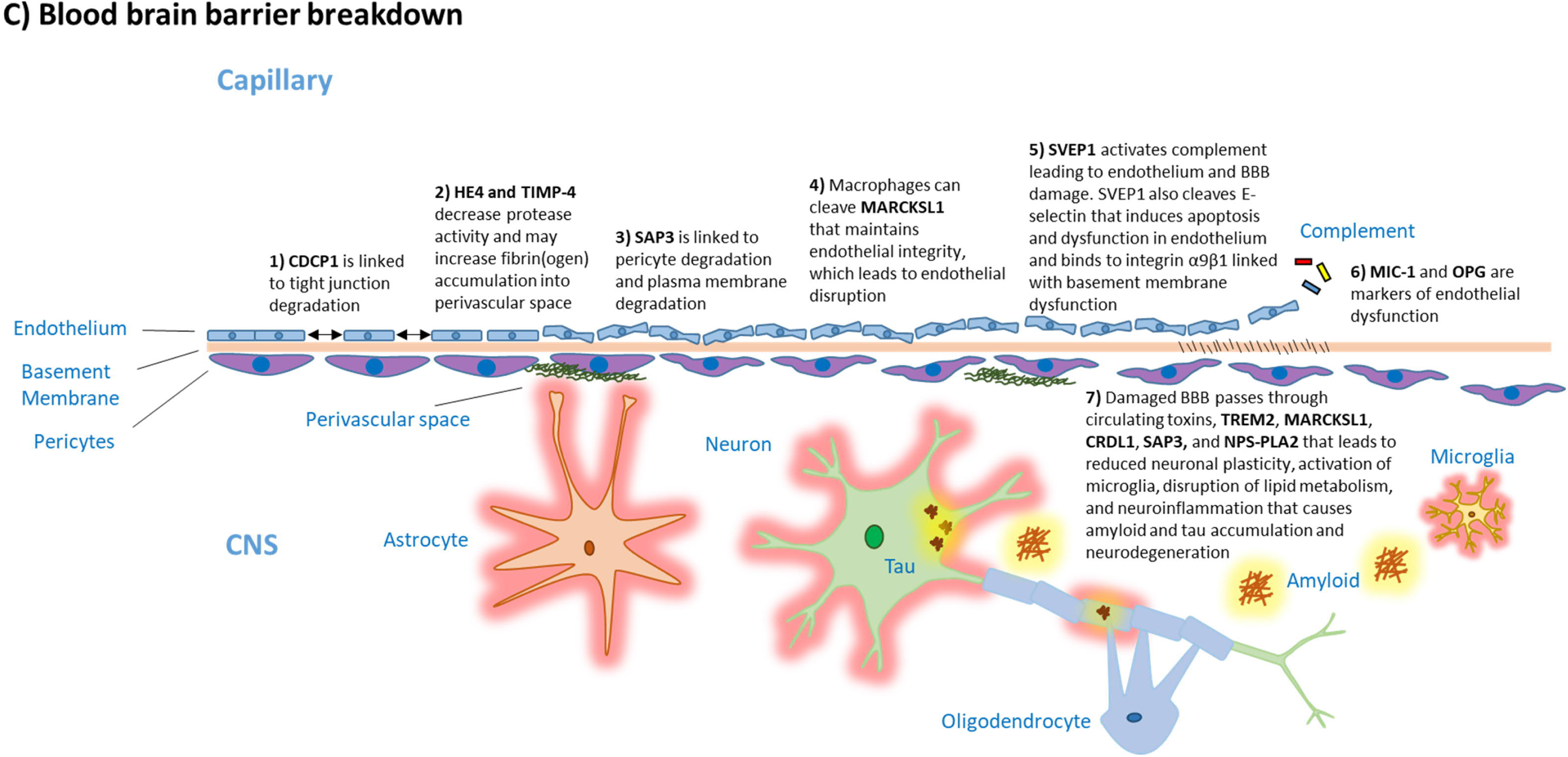
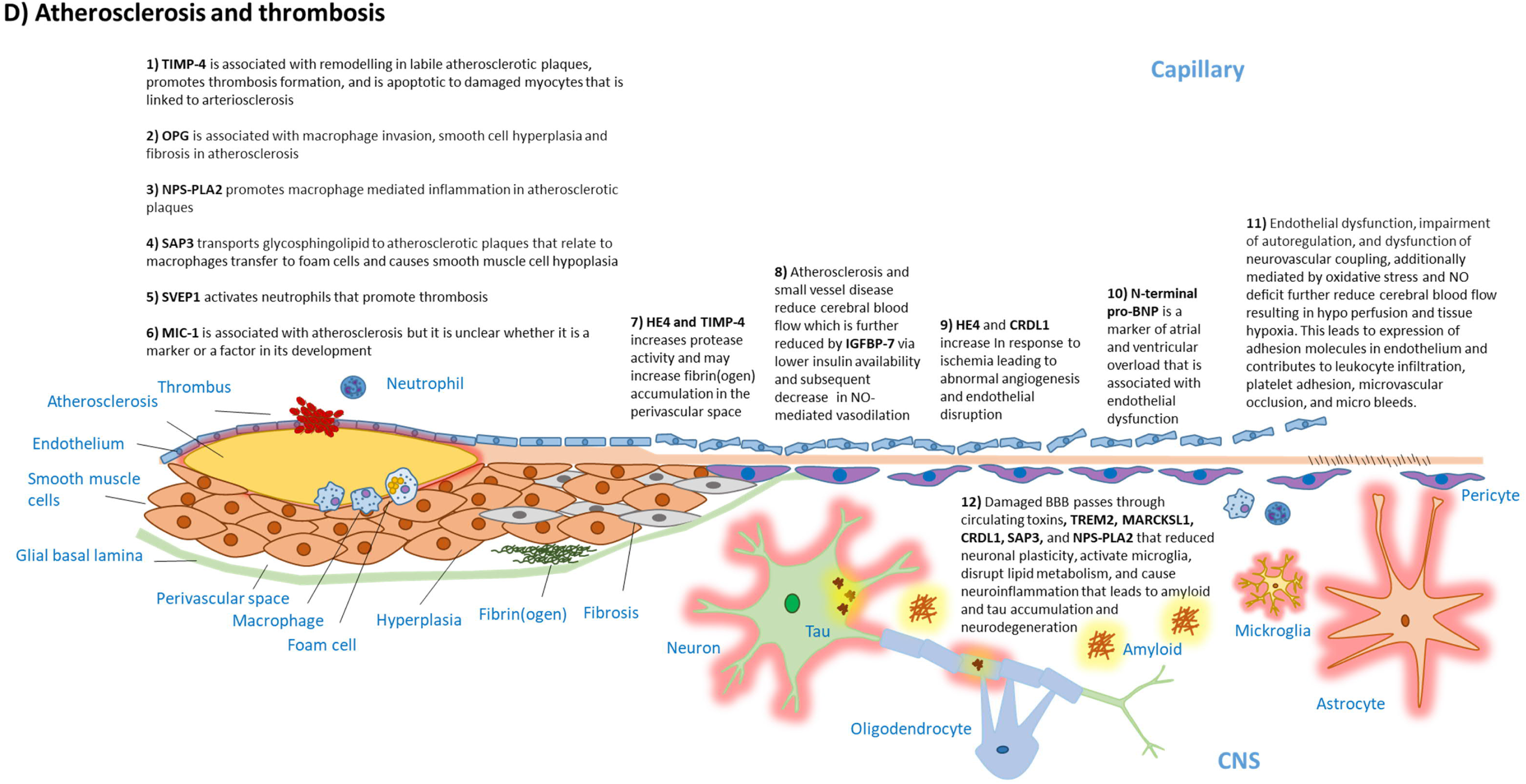
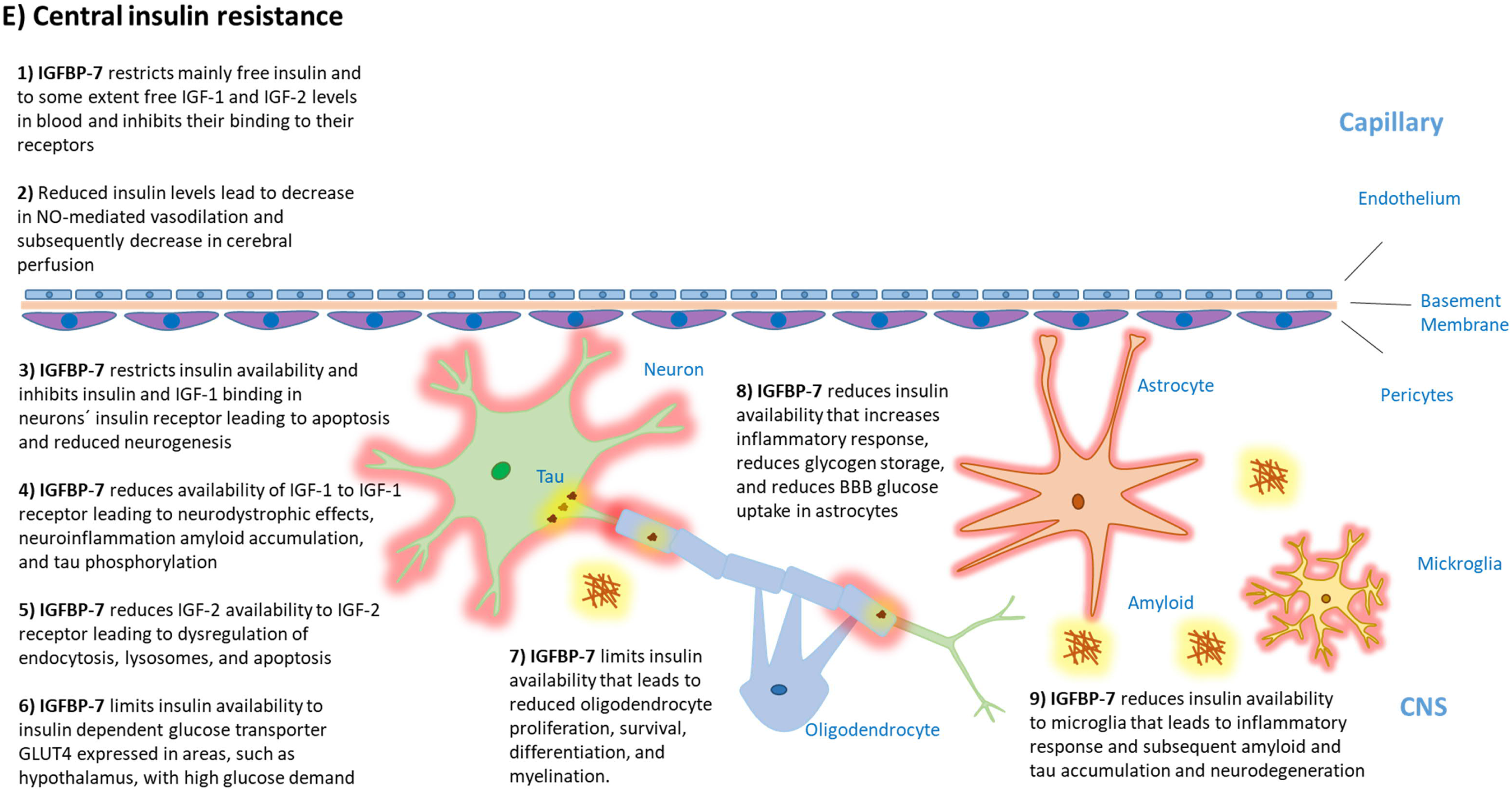
Dementia-related pathologies for the 15 proteins associated with cognitive decline rate and increased dementia incidence

CDCP1 is a further identified immune-system related protein. It is a ligand for CD6 on T-cells, which upregulates T-cell activation and infiltration, and increases synthesis of pro-inflammatory cytokines.^42^ According to multiple sclerosis mouse models, anti-CD-6 antibodies ameliorate, and CD6 and CDCP1 knock out protects from encephalomyelitis^42^ suggesting that higher levels of CDCP1 might predispose to central inflammation. SIGLEC-7 is an inhibitory receptor in NK-cells that recognizes “self” antigens and prevents NK cell attacks on healthy cells.^43^ In uncontrolled inflammation, SIGLEC-7 is cleaved and released into the plasma.^44, 45^ Thus elevated plasma SIGLEC-7 may reflect NK-cell activation towards healthy tissues, a potential risk factor for BBB dysfunction and CNS damage.

HE4 and RNAS6, in turn, are involved in antimicrobial responses to pathogens. HE4 is a broad-spectrum protease inhibitor^46^ protecting against proteolytic microbial virulence. RNAS6 belongs to antimicrobial proteins and peptides (AMPs) that are secreted to blood in response to pathogens^47, 48^ after infection has breached epithelium.^47, 48^ The influence of direct antimicrobial responses in dementia pathogenesis is supported by the antimicrobial protection hypothesis,^49^ which suggests that amyloid β can act as an AMP and protect against pathogens in the CNS.^49^ However, prolonged activation of this process leads to amyloid deposition.

In combination, these findings support the hypotheses that inflammation and pathogens can cause chronic hyper-activation of innate and adaptive immunity, which then contributes to the development of Alzheimer′s disease and vascular dementia.^41, 49, 50^

Proteins SAP3, TIMP-4, MIC-1, and OPG have been found to be related to BBB degeneration (**Figure 4C**). Elevated SAP3 levels have been observed in the cerebrospinal fluid of Alzheimer’s and Parkinson’s disease patients^19, 51^ and are linked with pericyte degeneration in the retina,^52^ leading to retinopathy as a complication of diabetes.^53^ As pericyte degeneration is also observed in BBB breakdown, both mechanisms could play a role in development of dementia. TIMP-4 regulates the activity of multiple metalloproteinases^54^ that can predispose to BBB degeneration,^6^ but it is unclear whether elevated TIMP-4 levels are indicators of BBB-degrading processes or damage to the BBB by fibrinogen accumulation. Increased MIC-1 and OPG are also thought to reflect endothelial dysfunction.^55-57^ CDCP1 is linked with tight junction degeneration^58-60^ and HE4 is a broad spectrum proteinase inhibitor,^46^ similar to TIMP-4. MARCKSL1 participates in maintaining endothelia integrity and when cleaved, its elevated plasma levels may reflect endothelial damage.^61, 62^ SVEP1 can cleave E-selectin that induces apoptosis and dysfunction in endothelium and binds to integrin α9β1 linked with basement membrane dysfunction.^63^

Damaged BBB passes circulating TREM2, MARCKSL1, CRDL1, SAP3 and NPS-PLA2 into the CNS contributing to reduced neuronal plasticity, activation of microglia, disruptions in lipid metabolism, and increased neuroinflammation, amyloid and tau accumulation, and neurodegeneration.^7, 30, 31, 64-67^ These findings support the hypothesis that BBB degradation with subsequent passage of toxins and proteins into CNS may play an important aetiological role in dementia.^6^ We are not aware of previous studies showing that circulating TREM2 already in middle adulthood is associated with increased dementia risk at older ages.

Seven previously identified and three novel proteins are likely to play a role in vascular pathologies (**Figure 4D**). TIMP-4, SAP3, NPS-PLA2, OPG, and MIC-1 have been linked to dementia and atherosclerosis, remodelling of atherosclerotic plaques, platelet aggregation and thrombosis, and leukocyte invasion into atherosclerotic plaques.^54, 55, 57, 68-73^ N-terminal pro-BNP is known to be elevated in response to atrial and ventricular overload^74^ and in plasma and the CNS it can inhibit the activity of neprilysin, an enzyme that reduces amyloid β levels.^75^ Elevated N-terminal pro-BNP levels are also linked with increased risk of vascular and Alzheimer’s dementia.^28, 29, 76, 77^ Of the proteins not previously linked to dementia, SVEP1 is known to promote thrombosis,^78, 79^ and HE4 and CRDL1 participate in angiogenesis in response to ischemia^80-82^ and might contribute to the abnormal angiogenesis observed in vascular and Alzheimer’s dementia.^6, 83^ These results provide a variety of mechanisms that could predispose to cerebrovascular dysfunction in dementias.

IGFBP-7 is a protein with high affinity to insulin, and lower affinity to growth factors IGF-1 and IGF-2.^84, 85^ It is related to reduced central blood flow by restricting free insulin^85^ in blood which leads to decreased insulin-NO-mediated vasodilation that can predispose to ischemia (**Figures 4D and E)**. IGFBP-7 also prevents insulin from binding to its receptor.^85^ All these properties are linked to reduced insulin responsiveness indicating that IGFBP-7 may be one of the factors behind central insulin resistance linked with dementia,^86^ a hypothesis supported by a case-control study showing elevated IGFBP-7 levels in the urine of Alzheimer’s dementia patients.^20^

Our findings are of potential importance to drug repurposing and research on preventive medication for dementia. Varespladib inhibits the arachidonic acid pathway in inflammation by inhibiting NPS-PLA2 activity, and may counteract inflammation that predisposes to dementia.^87^ Similarly, itolizumab prevents CDCP1 from binding to CD6, leading to downregulation in T-cell mediated inflammation.^88^ Randomized control trials on intranasal insulin have shown promising results in improving amyloid and tau profiles and cognition in individuals with cognitive decline or Alzheimer’s dementia.^86^ Whether more tailored medication that targets IGFBP-7 can be used to ameliorate central insulin resistance should be investigated. Glucose lowering and statin medications linked with reduced inflammation in diabetes are also associated with reduced OPG levels;^56, 89, 90^ further research should determine whether OPG plays a role in the disease process leading to dementia. Currently available antihypertensive medications can be used to reduce plasma N-terminal pro-BNP levels,^74^ but whether a drug that directly targets N-terminal pro-BNP is beneficial for cognitive decline and dementia needs to be tested.

This study’s strengths include its large sample size, simultaneous assessment of thousands of proteins, replication of main findings in an independent study population, and the early baseline that allowed assessment of proteins two decades before dementia diagnosis when no changes in plasma amyloid or tau markers were present. Only three dementia cases occurred within the first 10 years of follow-up suggesting low risk of reverse causation. However, observational evidence cannot determine causal associations and therefore our findings should be considered as hypothesis generating. Other limitations include the lack of repeated protein measurements to assess whether changes in protein levels over time are associated with dementia risk; the use of semi-quantitative protein data that cannot determine clinically useful concentrations; and the lack of information on dementia subtypes based on brain imaging or cerebrospinal fluid biomarkers.

In conclusion, using large-scale testing of plasma proteins as long-term risk factors for cognitive decline and dementia, we have provided strong support for the hypothesis that early systemic processes may drive development of dementias. The protein-dementia associations identified and replicated were plausible, involving innate and adaptive immunity, BBB dysfunction, vascular pathology, and central insulin resistance that all can contribute to amyloid and tau deposition, or pathologies that characterise vascular dementia. Our findings point to proteins that may be potential biomarkers and drug targets for dementia, the first crucial step to discover disease-modifying medications that can expand the field beyond amyloid and tau.

## Methods

### Study design and participants

We used the Whitehall II study as our discovery cohort and the ARIC study as the replication cohort (**Figure 1**). In 1985-1988, all civil servants aged 35 to 55 years based in 20 departments in London, UK, were invited to participate in the Whitehall II cohort study, and 73% (n=10 308) agreed.^91^ Blood samples for proteomic analyses were collected from a random subsample of 2274 dementia-free individuals in 1997-99.^92^ Cognitive performance measurements were conducted at this and four subsequent clinical examinations in 2002-2004, 2007-2009, 2012-13, and 2015-2016. Follow-up started from 1997-99 and ended at death, dementia, or October 2019.

ARIC^93^ is a community-based cohort study of 15,792 participants from four US communities: Washington County, Maryland; Forsyth County, North Carolina; North-western suburbs of Minneapolis, Minnesota; and Jackson, Mississippi. Participants were aged 44 to 66 years at baseline in 1987-1989. Blood samples for protein analysis were drawn in 1993-1995 and analysed for 11 395 individuals free of dementia. Incident dementia cases were identified during the 1996-1998, 2011-2013 and 2016-2017 clinical examinations and using data from informant interviews, hospital records and death certificates.

### Assessment of plasma proteins

In Whitehall II and ARIC, proteins were analysed using the SomaScan version 4 assay. Earlier studies describe performance of the SomaScan assay and the modified aptamer-binding in detail.^94-96^ In brief, the SomaScan Assay is a multiplexed aptamer-based immunoassay that uses a library of DNA molecules designed to identify specific proteins. The aptamers bind to the target protein in plasma samples and the unspecific fast off rate proteins are washed away. The slow off rate proteins are labelled with biotin and the aptamer-protein complexes incubated in a buffer containing unlabelled polyanionic competitor. This step is analogous to the effect of adding a second antibody to increase specificity in a conventional immunoassay. The aptamer-protein complexes are then recaptured with streptavidin and again washed to remove non-specific aptamer-protein complexes. The aptamers are then released from the proteins, hybridized to complementary sequences on a DNA microarray chip, and quantified by fluorescence. The fluorescence intensity reflects the concentration of each protein. Median intra- and inter-assay coefficients of variation are ∼5% and assay sensitivity is comparable to that of typical immunoassays, with a median lower limit of detection in the femtomolar range.^95^ Specificity of the modified aptamer reagents is good and has been tested in several ways.^92^

### Cognitive testing

The Whitehall II cognitive test battery covered four domains: executive function, memory, phonemic-, and semantic fluency (**Supplementary Methods**). Executive function was assessed with the Alice Heim 4-I test, memory with a 20-word free recall test and phonemic and semantic fluency with word and animal name recall tests.^97, 98^ To minimize measurement error inherent in individual tests, we created a global cognitive score by standardizing each test domain measured in follow-up visits to the baseline score, and then standardizing the summary scores from each phase to the baseline summary score.^99^ This allowed construction of individual slopes of decline in cognition.

### Dementia follow-up

Whitehall II study participants are linked to the National Health Services (NHS) Hospital Episode Statistics (HES) database, and the British National mortality register using individual NHS identification numbers for linkage.^100^ The NHS provides nearly complete comprehensive health care coverage for all individuals legally resident in the UK. Incident dementia was defined by the WHO International Classification of Diseases, version 10 (ICD-10) codes F00, F01, F03, G30, and G31 and ICD-9 codes 290.0-290.4, 331.0-331.2, 331.82, and 331.9. We also conducted informant interviews and checked participants’ medications at each screening for dementia-related medication. Sensitivity and specificity of dementia assessment based on the HES data is 0.78 and 0.92.^101^

In ARIC, incident dementias were primarily identified using comprehensive clinical and neuropsychological examinations and informant interviews from visits 5 (2011-2013) and 6 (2016-2017).^102, 103^ Using all the data available, suspected cases were adjudicated by a committee of clinicians. In between visits, participants were contacted annually via telephone and administered a brief cognitive screener, and their caregivers, a questionnaire. This information, supplemented by surveillance of dementia-related hospital discharge and death certificate codes, was used to estimate the dementia onset date.

### Measurement of baseline covariates

In the Whitehall II study, standard self-administered questionnaires provided data on age, sex, ethnicity, socioeconomic status, education, alcohol consumption, and smoking. Experienced clinical nurses measured BMI and systolic blood pressure (SBP) and took blood samples for lipid and glucose measurements.^91^ Using DNA extracted from whole blood, a standard PCR assay determined *APOE* genotype using the salting out method.^104, 105^ Two independent observers read the genotype blind and any discrepancies were resolved by repeating the PCR analysis. In ARIC, baseline covariates included age, sex, and a combined race-study centre variable.

### Statistical analysis

We used Spearman correlation to test protein-protein correlations. In the Whitehall II study, individual cognitive measurements were plotted and group means derived with local linear smooth plots for descriptive pourposes.^106^ The proteins were transformed to normal distribution by inverse rank-based normal transformation. We used age and sex adjusted linear regression to study associations between each protein and the cognitive decline slopes, derived with mixed-effects linear regression, and formed a Manhattan plot based on –log10(p-values). The assumptions of linear regression were assessed by plotting the residuals in residuals versus fitted, normal Q-Q, scale-location, and residual versus leverage plots.^106^ We used false discovery rate correction (FDR) of 5% to select proteins for further analyses; this translated to p-value cut-off of 0.002. The proteins that survived FDR correction were analysed in age, sex, and ethnicity adjusted Cox regression models^107^ with incident dementia as the outcome and p-value=0.05 as the cut-off for statistical significance. All the p-values were two-sided. The proportionality assumption in all Cox models was assessed with Schoenfeld residuals and with log-log plots.^107^ In sensitivity analyses Cox regression was adjusted for age, sex, ethnicity, SBP, antihypertensive medication, BMI, alcohol consumption, smoking, diabetes, socio-economic, and *APOE* status. We also studied the effect of competing risk of death with the Fine and Gray model.^108^ To examine reproducibility, proteins that were robustly associated with cognitive decline and dementia in the Whitehall II study were also analysed in the ARIC study. These Cox models with dementia as outcome were adjusted for age, sex and race-study centre.

We searched the Genotype-Tissue Expression (GTEx),^109^ Human Protein Atlas,^110, 111^ Database for Annotation, Visualization and Integrated Discovery (DAVID),^112, 113^ Uniprot,^114^ and ChEMBL^115^ databases to characterize protein expression profiles, their cellular localization, and drugs that can target them. We used statistical software R (3.6.0) and Stata (version 16.1 MP; Stata Corp, College Station, TX, USA) for all analyses.

## Data Availability

Data, protocols, and other metadata of the Whitehall II and ARIC studies are available to the scientific community. Please refer to the data sharing policies of these studies. Pre-existing data access policies for Whitehall II and ARIC studies specify that research data requests can be submitted to each steering committee; these will be promptly reviewed for confidentiality or intellectual property restrictions and will not unreasonably be refused. Individual-level patient or protein data may further be restricted by consent, confidentiality, or privacy laws/considerations. These policies apply to both clinical and proteomic data.

## Contributors

MK, AH, AS-M, GL, EJB, NM and JVL generated the hypothesis and designed the study. JVL, NM, KS and HZ provided initial mechanistic interpretations. JVL, with MK, wrote the first draft of the report. JVL did the primary analyses, with support from NM, and performed literature searches. KW ran the replication analysis. All authors interpreted the data and critically commented and reviewed the report. JVL had full access to pseudonymised data from the Whitehall II study and KW had full access to the ARIC data.

## Code availability

This manuscript did not include any custom code and used only pre-existing statistical packages.

## Acknowledgments

The study was supported by the UK Medical Research Council (S011676, K013351, R024227), the National Institute on Aging (National Institutes of Health; R01AG056477 and R01AG062553), the British Heart Foundation (32334), the Academy of Finland (311492), NordForsk (75021), and SomaLogic, Inc. The Genotype-Tissue Expression (GTEx) Project was supported by the Common Fund of the Office of the Director of the National Institutes of Health, and by NCI, NHGRI, NHLBI, NIDA, NIMH, and NINDS. The data used for the analyses described in this manuscript were obtained from: Median gene-level TPM by tissue dataset from the GTEx Portal on 9/2/2020. JVL was supported by Academy of Finland (311492) and Helsinki Institute of Life Science (H970) and AS-M and AGT were supported by NordForsk (75021, the Nordic Research Programme on Health and Welfare), during the conduct of the study. ASM is supported by the National Institute on Aging, NIH (R01AG062553;R01AG056477). RCL was supported by Alzheimer’s Research UK grant ARUK-NAS2017A-1. HZ is a Wallenberg Scholar supported by grants from the Swedish Research Council (2018-02532), the European Research Council (681712), Swedish State Support for Clinical Research (ALFGBG-720931), the Alzheimer Drug Discovery Foundation, USA (201809-2016862), and the UK Dementia Research Institute at UCL. EJB was supported by British Heart Foundation (RG/16/11/32334) and UK Research and Innovation (ES/T014377/1). MK was supported by UK Medical Research Council (MR/S011676, MR/R024227), US National Institute on Aging (R01AG062553, R01AG056477), Academy of Finland (311492), Helsinki Institute of Life Science (H970), NordForsk (75021), and the Finnish Work Environment Fund (190424). The Atherosclerosis Risk in Communities study has been funded in whole or in part with Federal funds from the National Heart, Lung, and Blood Institute, National Institutes of Health, Department of Health and Human Services (contract numbers HHSN268201700001I, HHSN268201700002I, HHSN268201700003I, HHSN268201700004I and HHSN268201700005I), R01HL087641, R01HL086694; National Human Genome Research Institute contract U01HG004402; and National Institutes of Health contract HHSN268200625226C. Neurocognitive data is collected by U01 2U01HL096812, 2U01HL096814, 2U01HL096899, 2U01HL096902, 2U01HL096917 from the NIH (NHLBI, NINDS, NIA and NIDCD). Infrastructure was partly supported by Grant Number UL1RR025005, a component of the National Institutes of Health and NIH Roadmap for Medical Research. This study was also supported by contracts K23 AG064122 (Dr. Walker) and K24 AG052573 (Dr. Gottesman) from NIA. UCL and Johns Hopkins University have signed a collaboration agreement with SomaLogic, Inc to conduct SOMAscan of Whitehall and ARIC stored samples at no charge in exchange for the rights to analyse linked Whitehall and ARIC phenotype data.

## Supplement

### Supplementary Methods

#### Cognitive testing

The Whitehall II cognitive test battery covered four domains: executive function, memory, phonemic-, and semantic fluency. Executive function was assessed with the Alice Heim 4-I test, that comprises a series of 65 verbal and mathematical reasoning items of increasing difficulty.^97^ It tests inductive reasoning, measuring the ability to identify patterns and infer principles and rules, and has time limit of 10 minutes. A 20-word free recall test assessed memory. Participants were presented a list of one or two syllable words at two-second intervals and were then asked to recall in writing as many of the words as possible in any order with two minutes to do so. In assessment of phonemic and semantic fluency^98^ participants were asked to recall in writing as many words beginning with “s” (phonemic fluency) and as many animal names (semantic fluency) as they could. Time limit for this section was one minute per fluency area.

Based on these measures, we created a global cognitive score by first standardizing the distribution of each test domain measured in follow-up visits to the baseline score to create z-scores with mean 0 and SD 1. We then summed the domain specific scores at each phase and standardized the summary score to the baseline summary score; this approach minimizes measurement error inherent in individual tests.^99^ After dementia diagnosis, participants were rarely able to complete the cognitive tests and for this reason, their global test score was set to −3SD at phase that followed dementia diagnosis. Based on the global scores we derived a cognitive decline slope for each participant and used this as the outcome.

---

**Supplementary Table 1.** Hazard ratios between SD increase in amyloid, tau, and neurofilament related protein levels and cognitive decline. None of these survived false discovery rate (FDR) correction of 5%.

| Protein | Cognitive decline |  |
| --- | --- | --- |
|  | Beta | FDR p-value |
| **Amyloid, tau, and neurofilament related proteins** |  |  |
| Amyloid beta precursor like protein 1 | 0.02 | 0.65 |
| Amyloid beta precursor like protein 2 | 0.03 | 0.35 |
| Serum amyloid A1 | 0.05 | 0.19 |
| Serum amyloid A2 | 0.05 | 0.13 |
| Serum amyloid A4 | 0.02 | 0.56 |
| Amyloid P component, serum | 0.02 | 0.53 |
| Amyloid beta precursor protein | 0.01 | 0.67 |
| Amyloid beta precursor protein binding family B member 1 | 0.03 | 0.35 |
| Amyloid beta precursor protein binding family B member 2 | 0.05 | 0.12 |
| Amyloid beta precursor protein binding family B member 3 | 0.02 | 0.63 |
| Microtubule associated protein RP/EB family member 1 | -0.01 | 0.75 |
| Microtubule associated protein RP/EB family member 2 | -0.01 | 0.77 |
| Microtubule associated protein RP/EB family member 3 | -0.02 | 0.63 |
| Microtubule associated proteins 1 light chain 3 alpha | 0.03 | 0.38 |
| Microtubule associated proteins 1 light chain 3 beta | 0.01 | 0.82 |
| Microtubule associated protein tau | 0.03 | 0.36 |
| Regulator of microtubule dynamics protein 1 | 0.01 | 0.86 |
| Regulator of microtubule dynamics protein 3 | 0.02 | 0.59 |
| Janus kinase and microtubule-interacting protein 3 | 0.06 | 0.05 |
| Microtubule affinity regulating kinase 3 | 0.01 | 0.92 |
| Neurofilament light | 0.01 | 0.69 |
| Neurofilament heavy | 0.04 | 0.20 |

**Supplementary Table 2.**
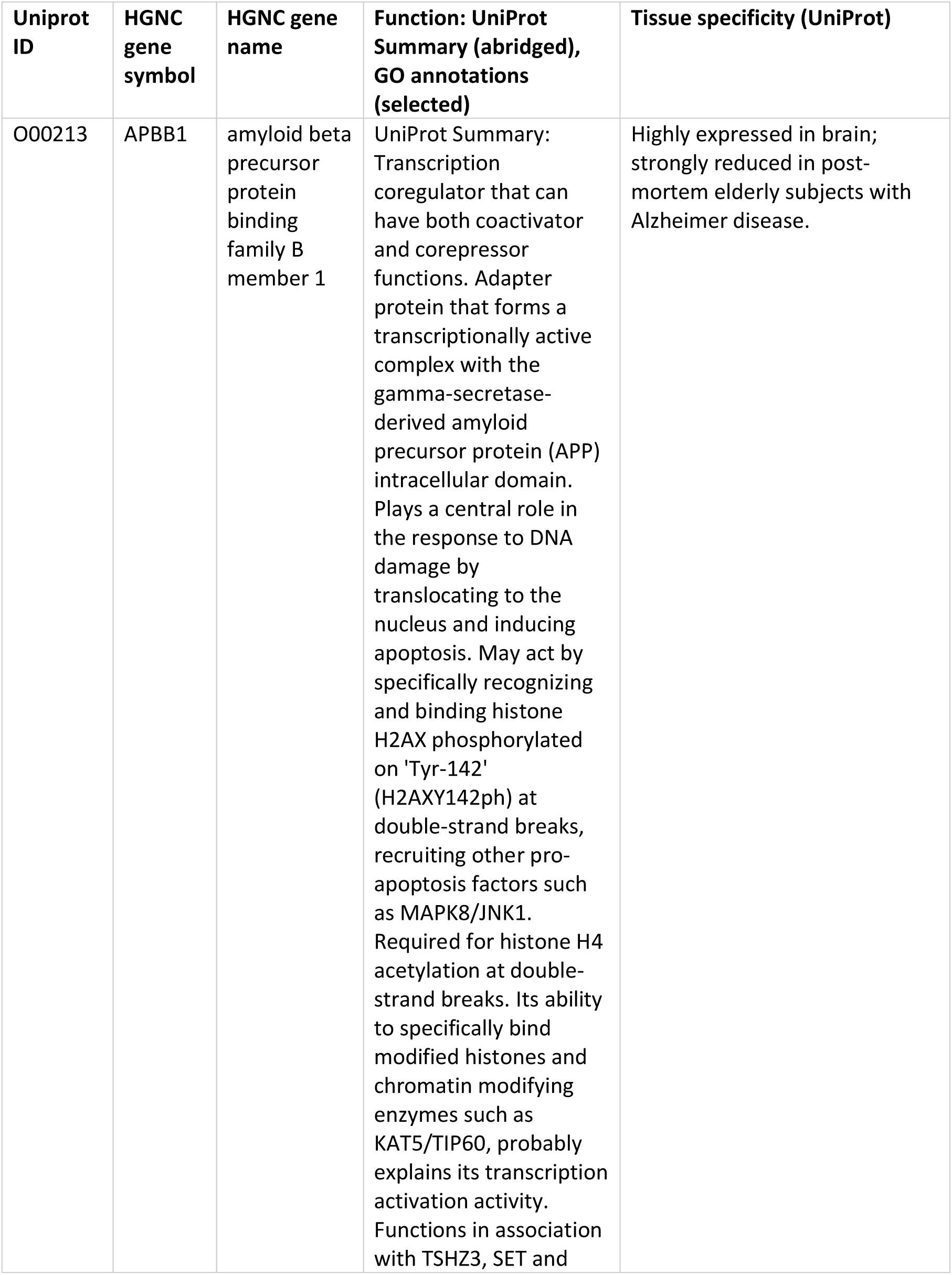
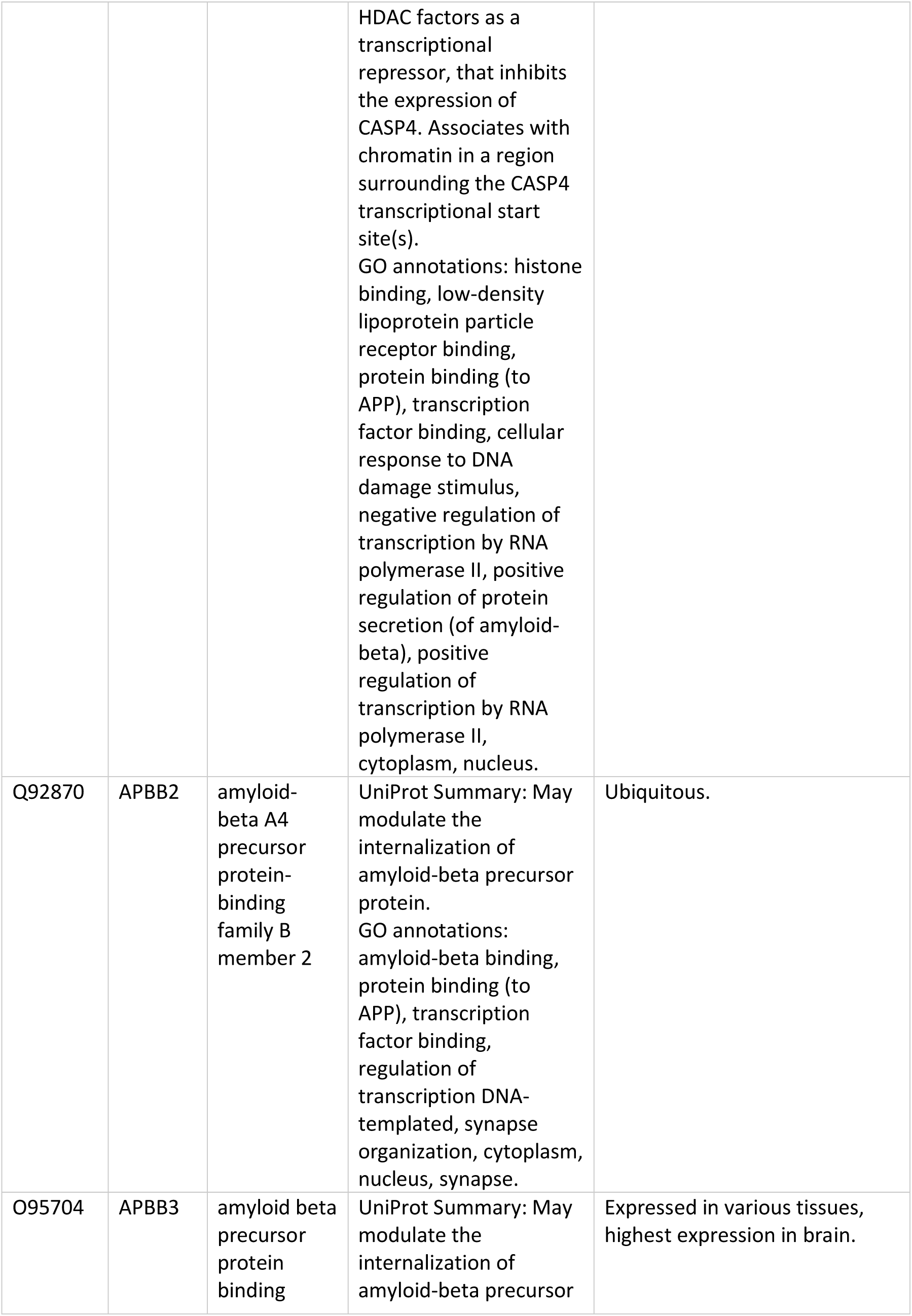
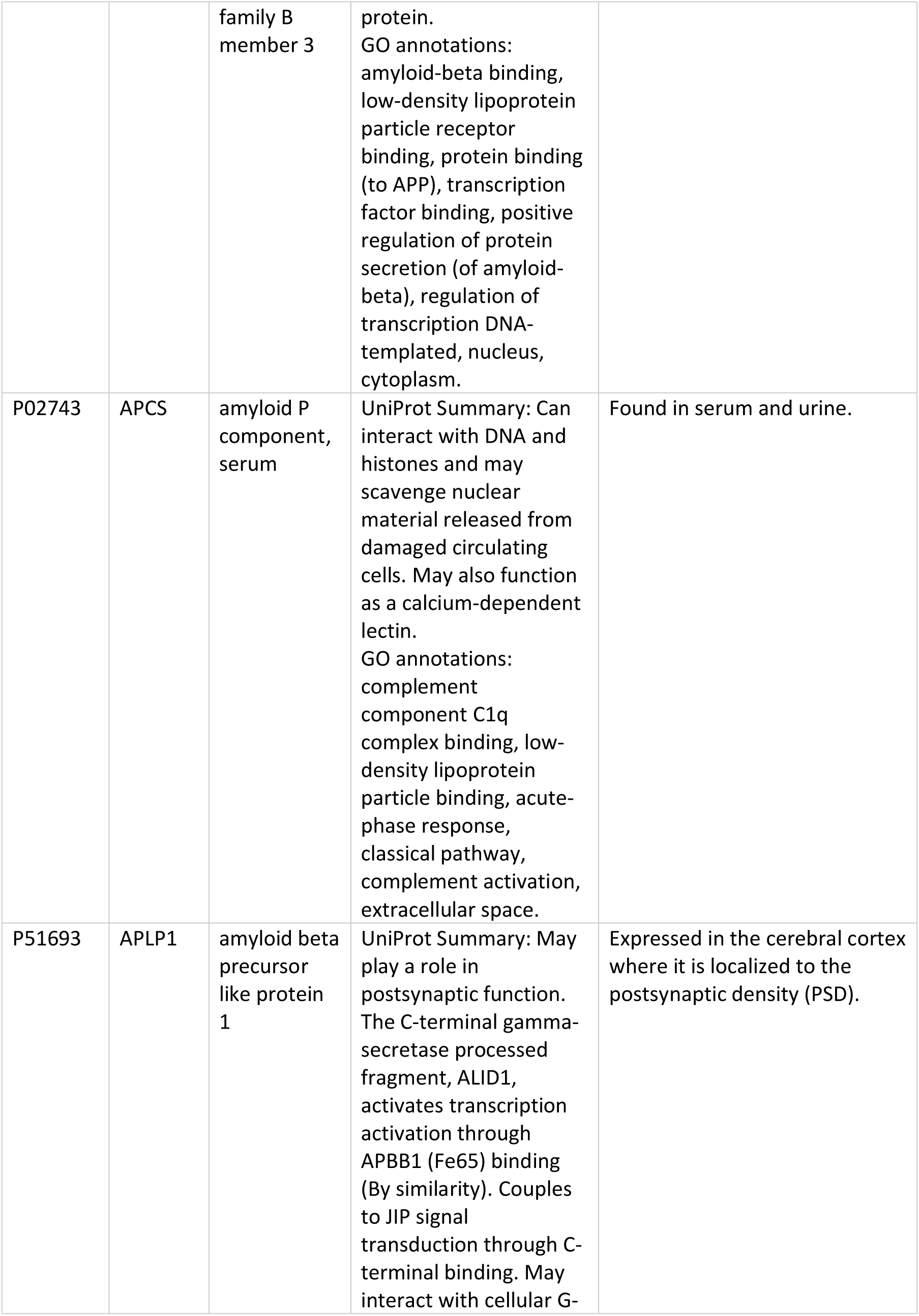
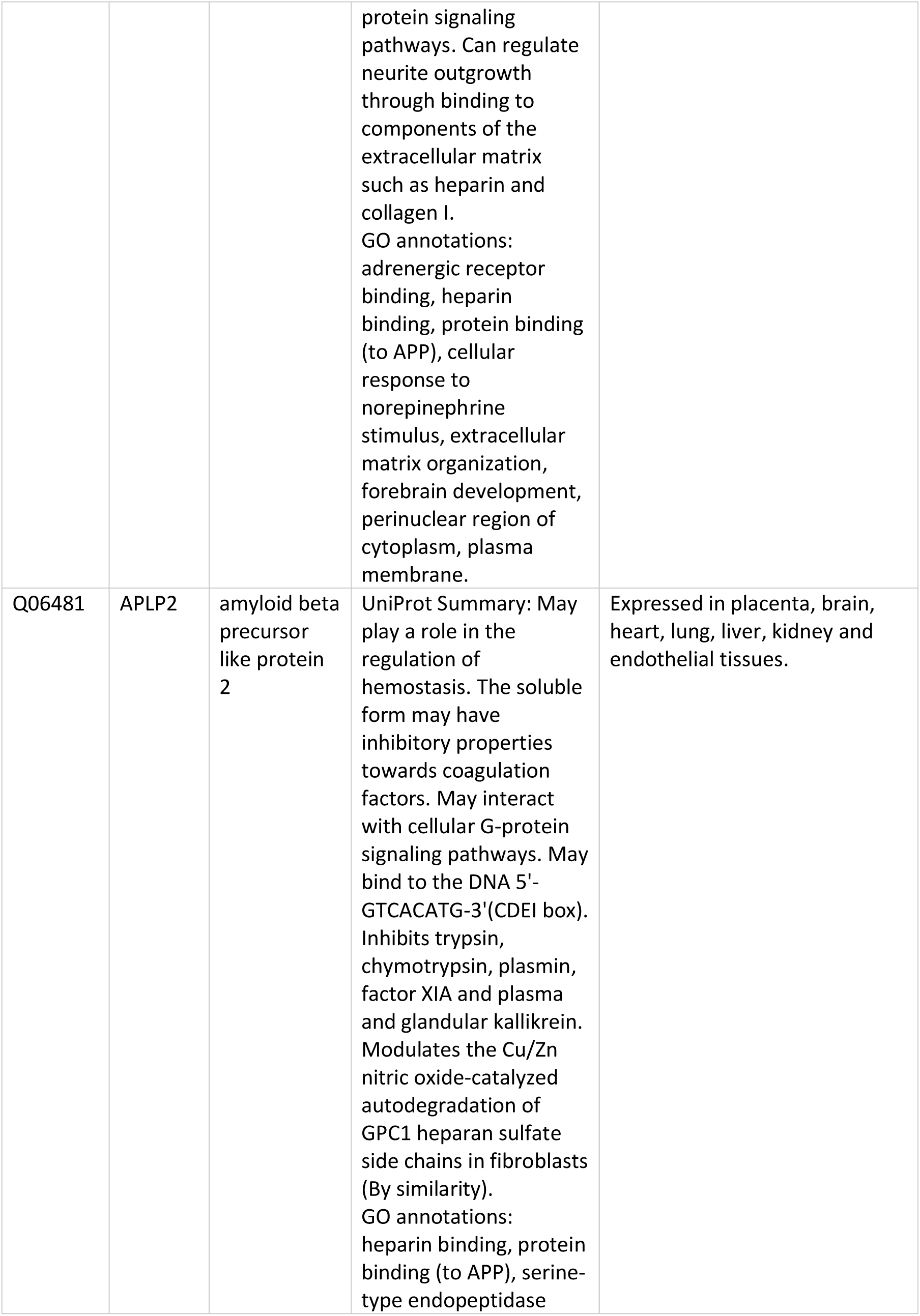
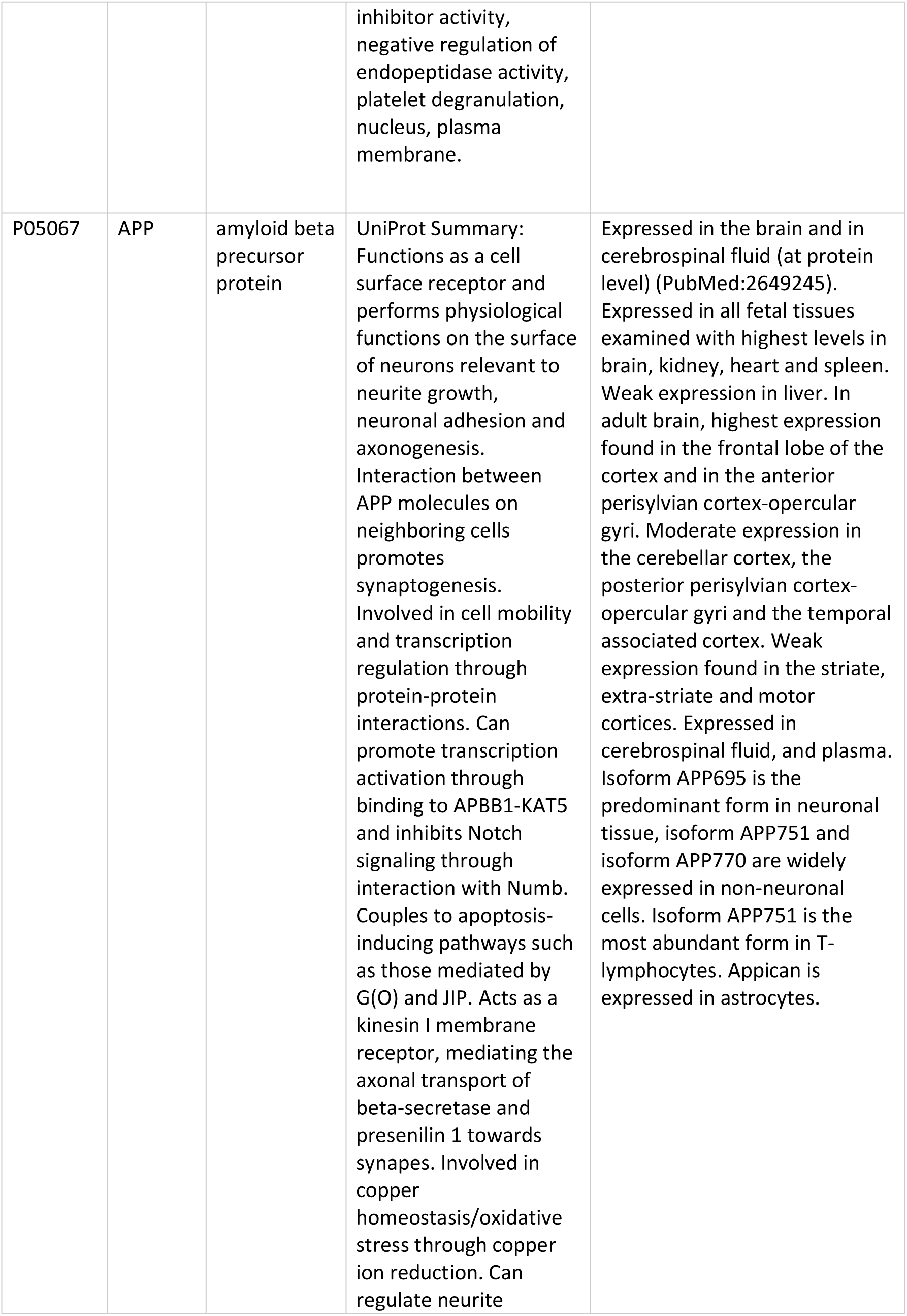
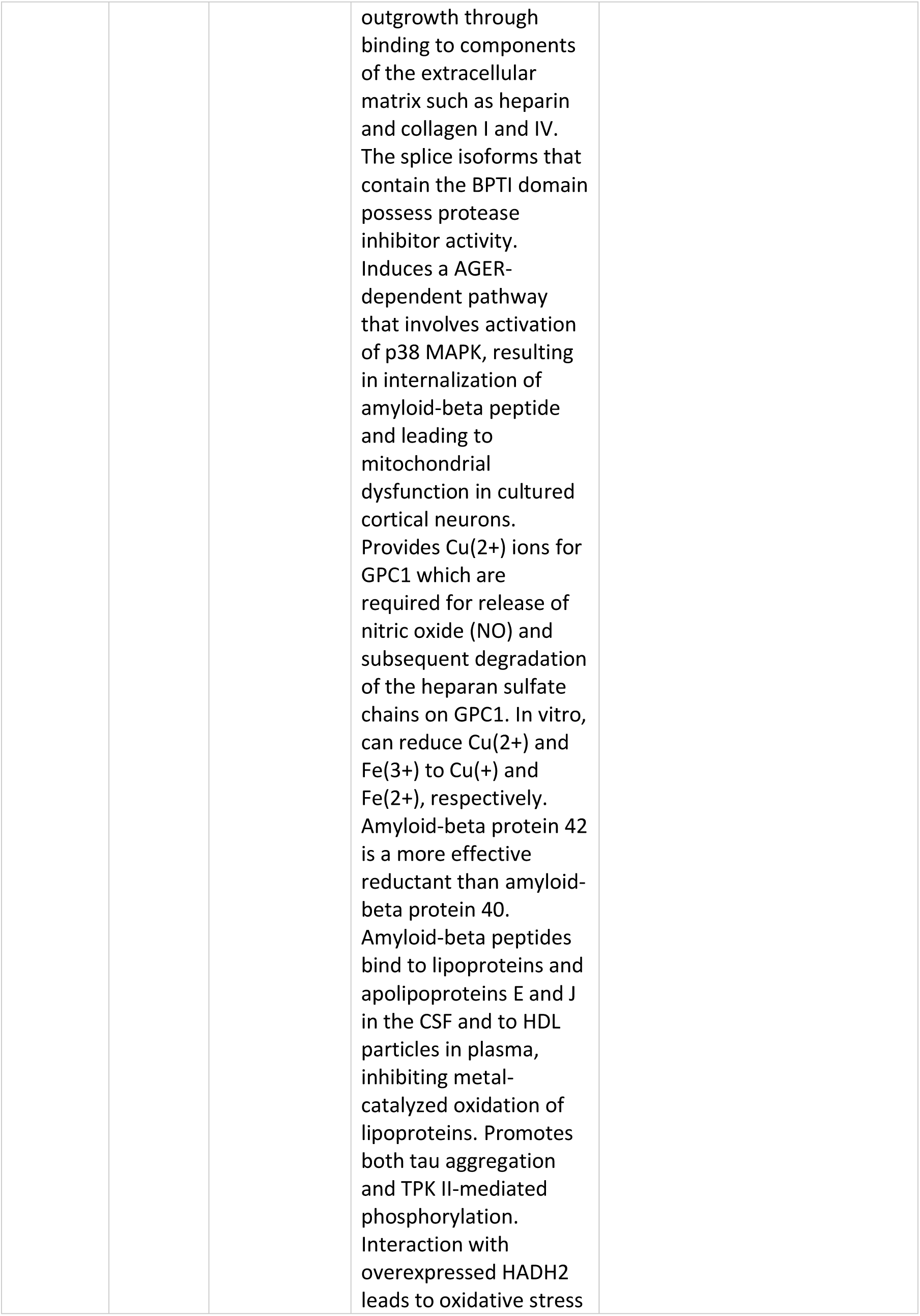
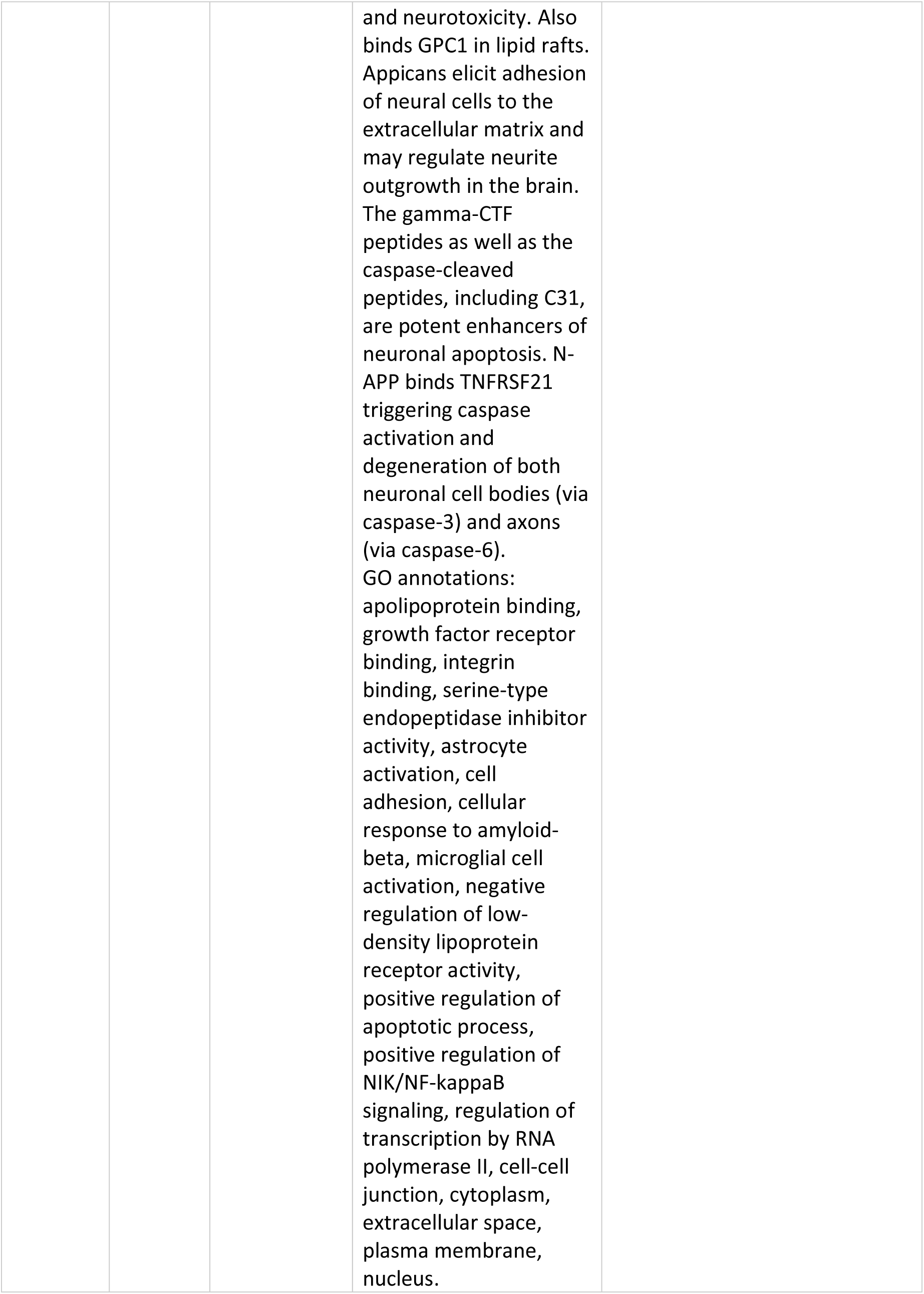
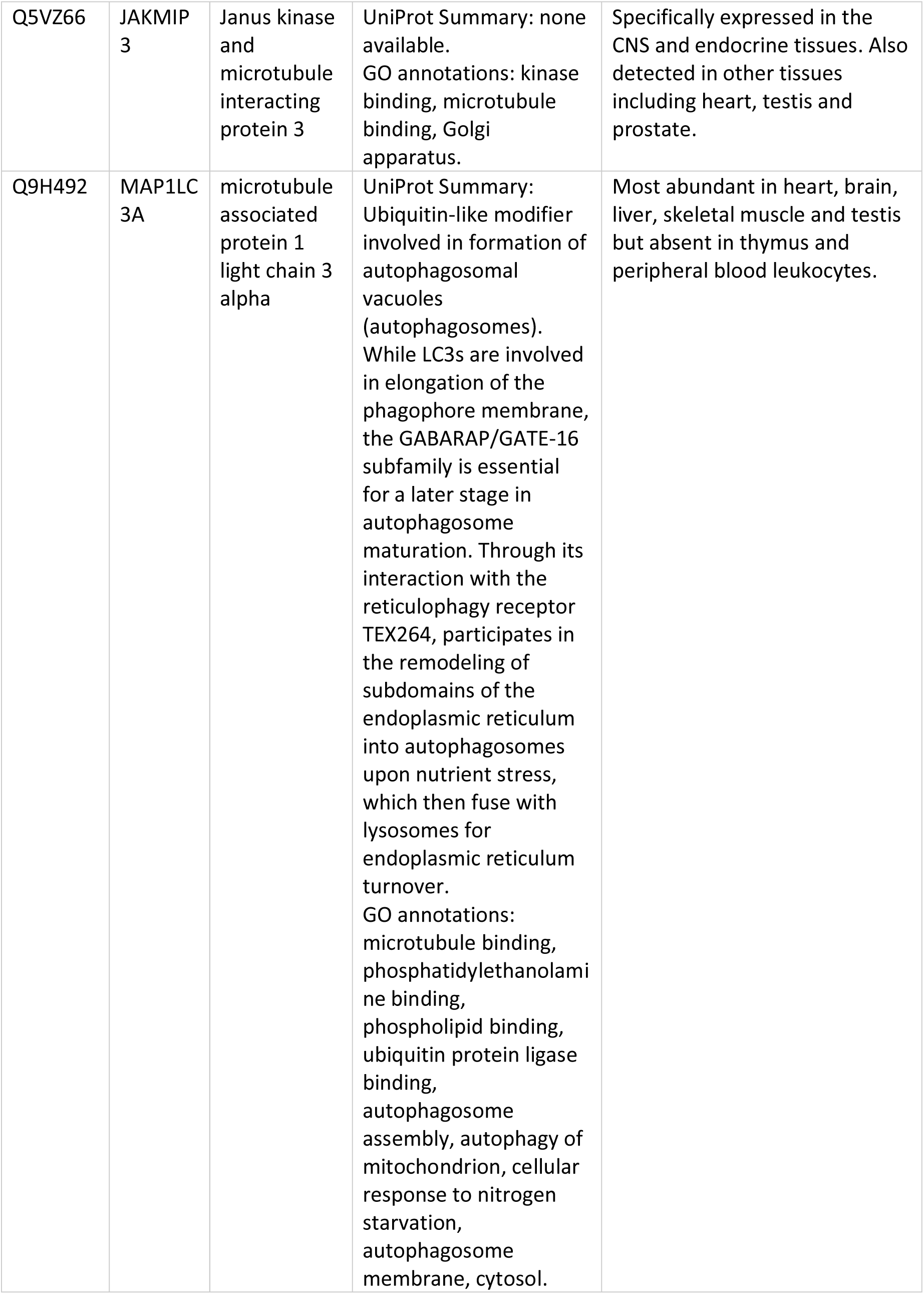
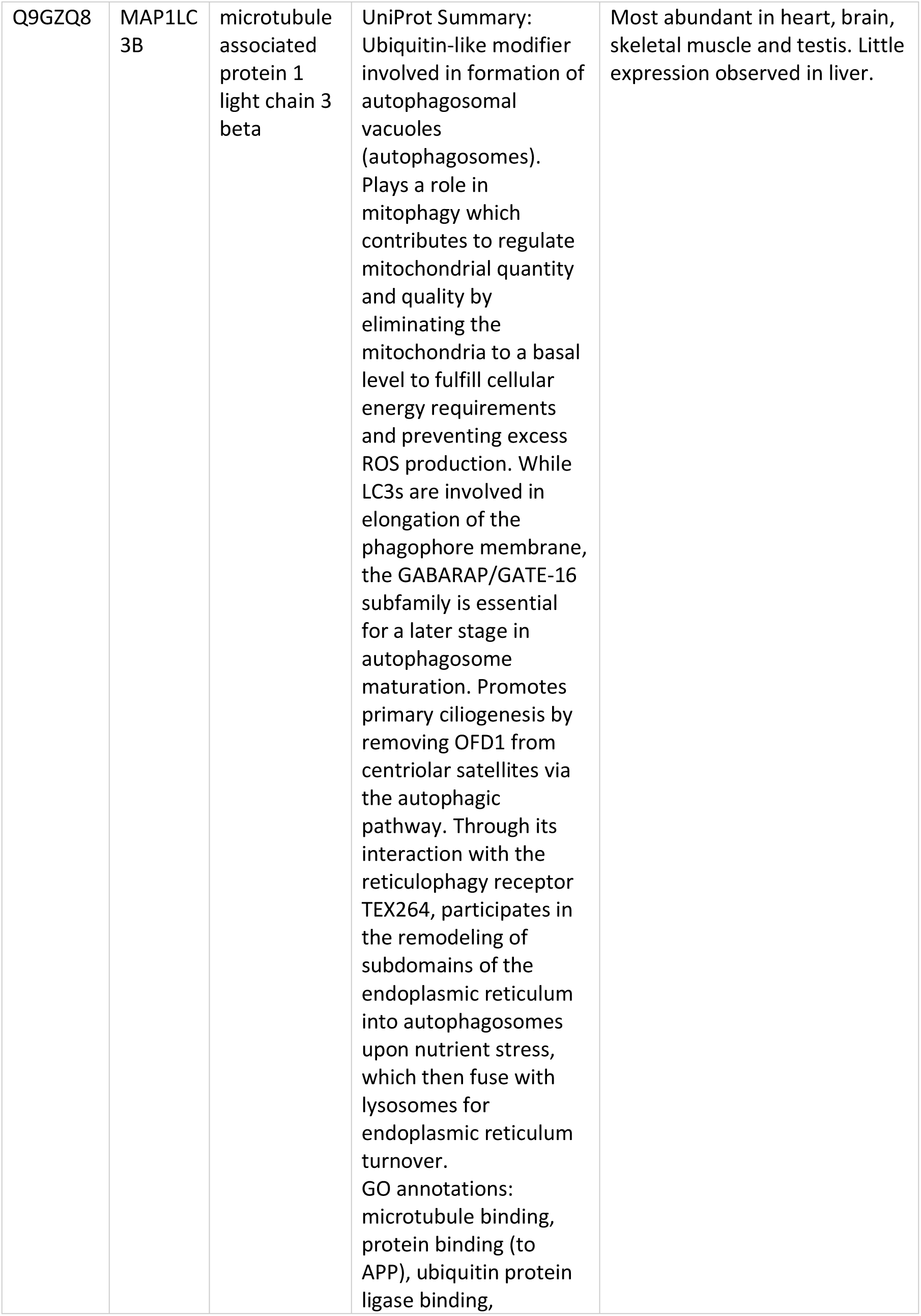
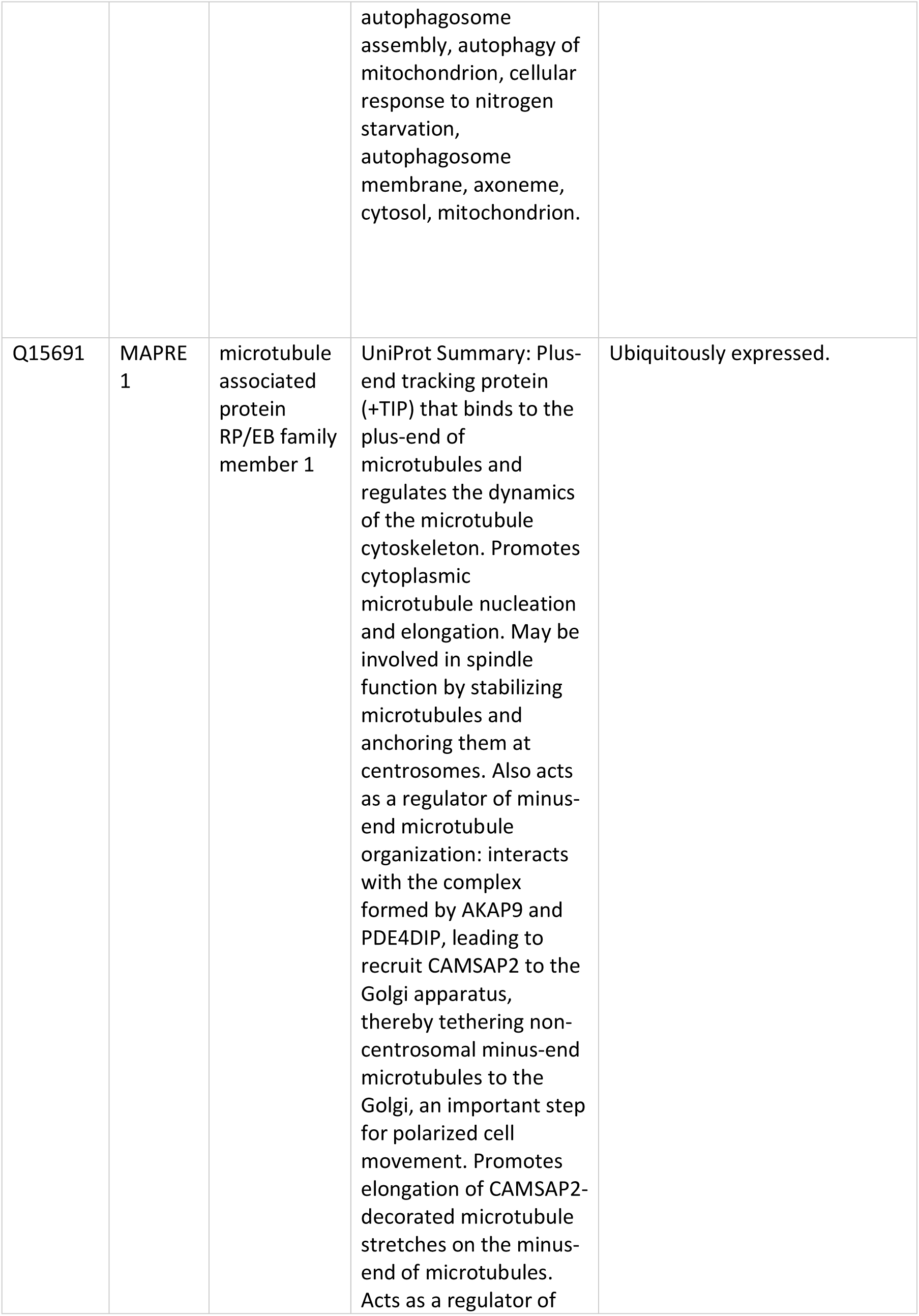
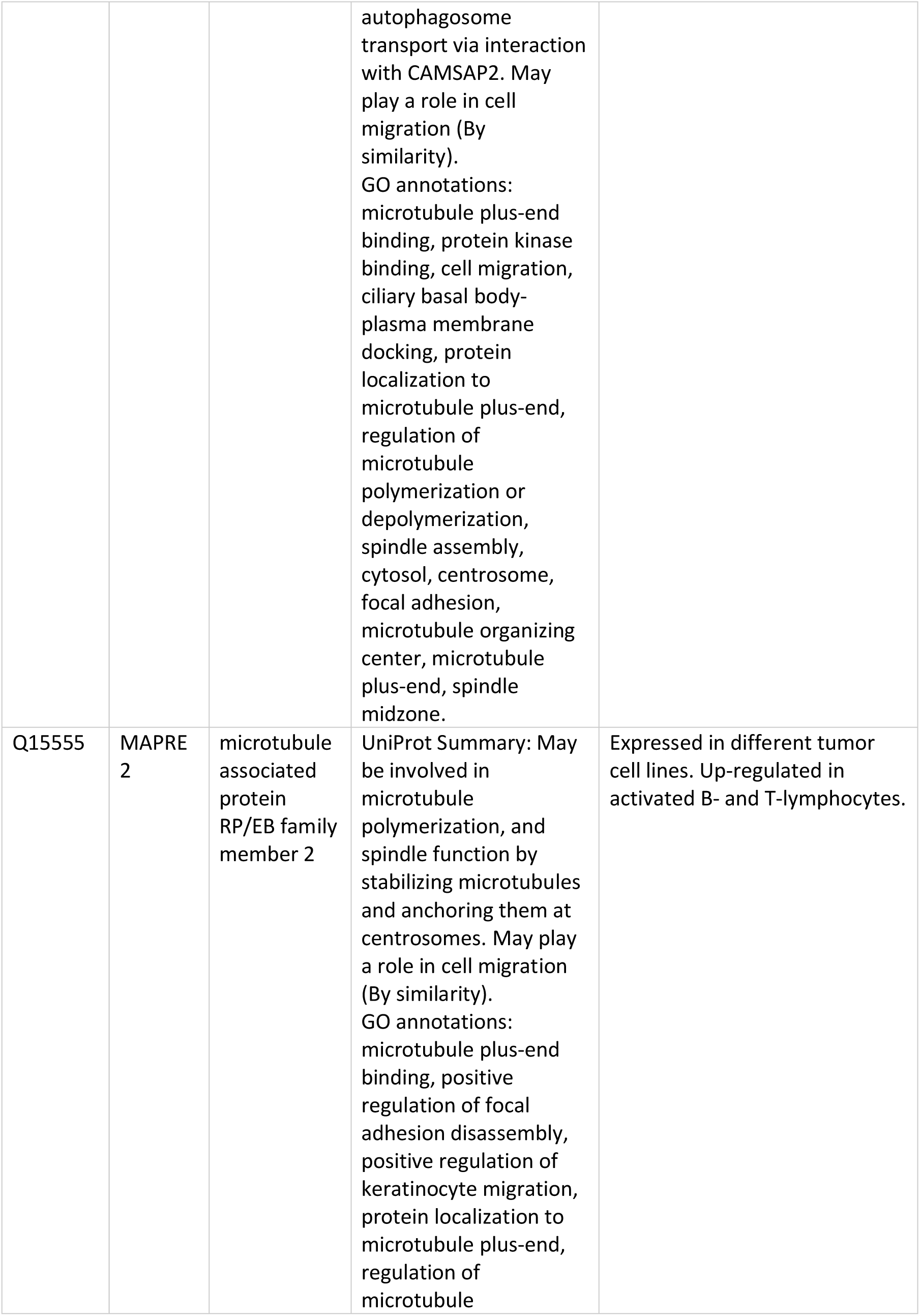
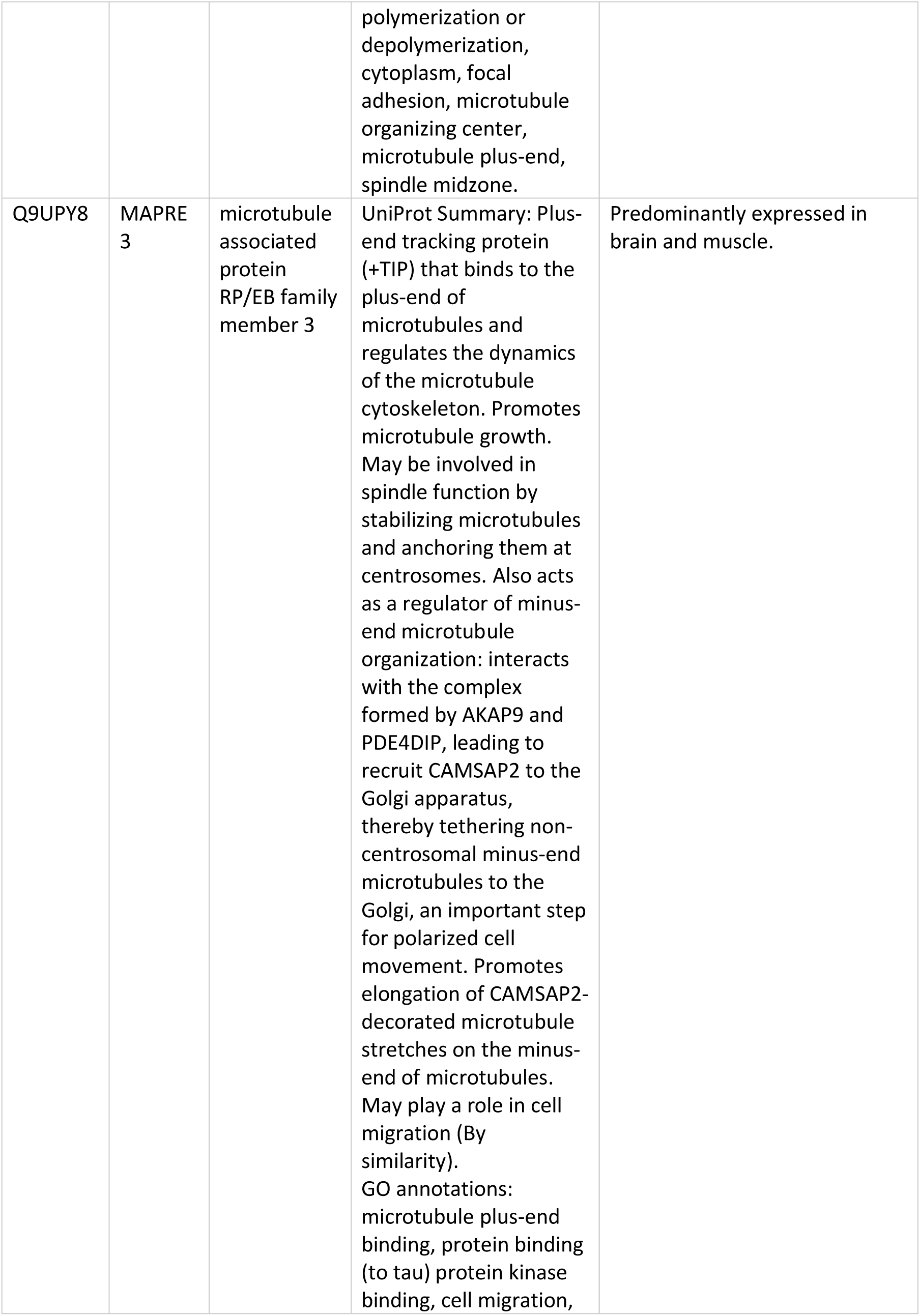
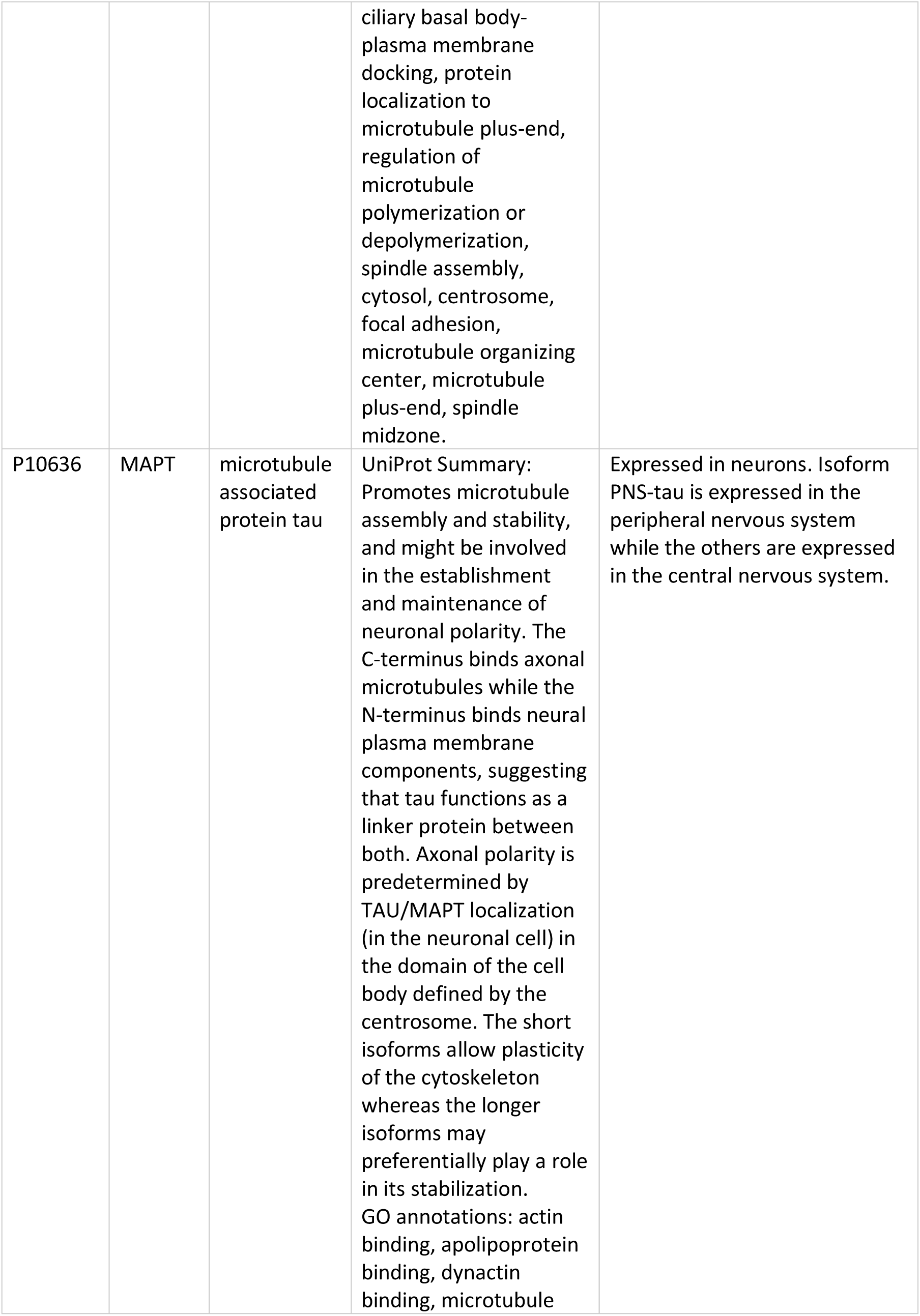
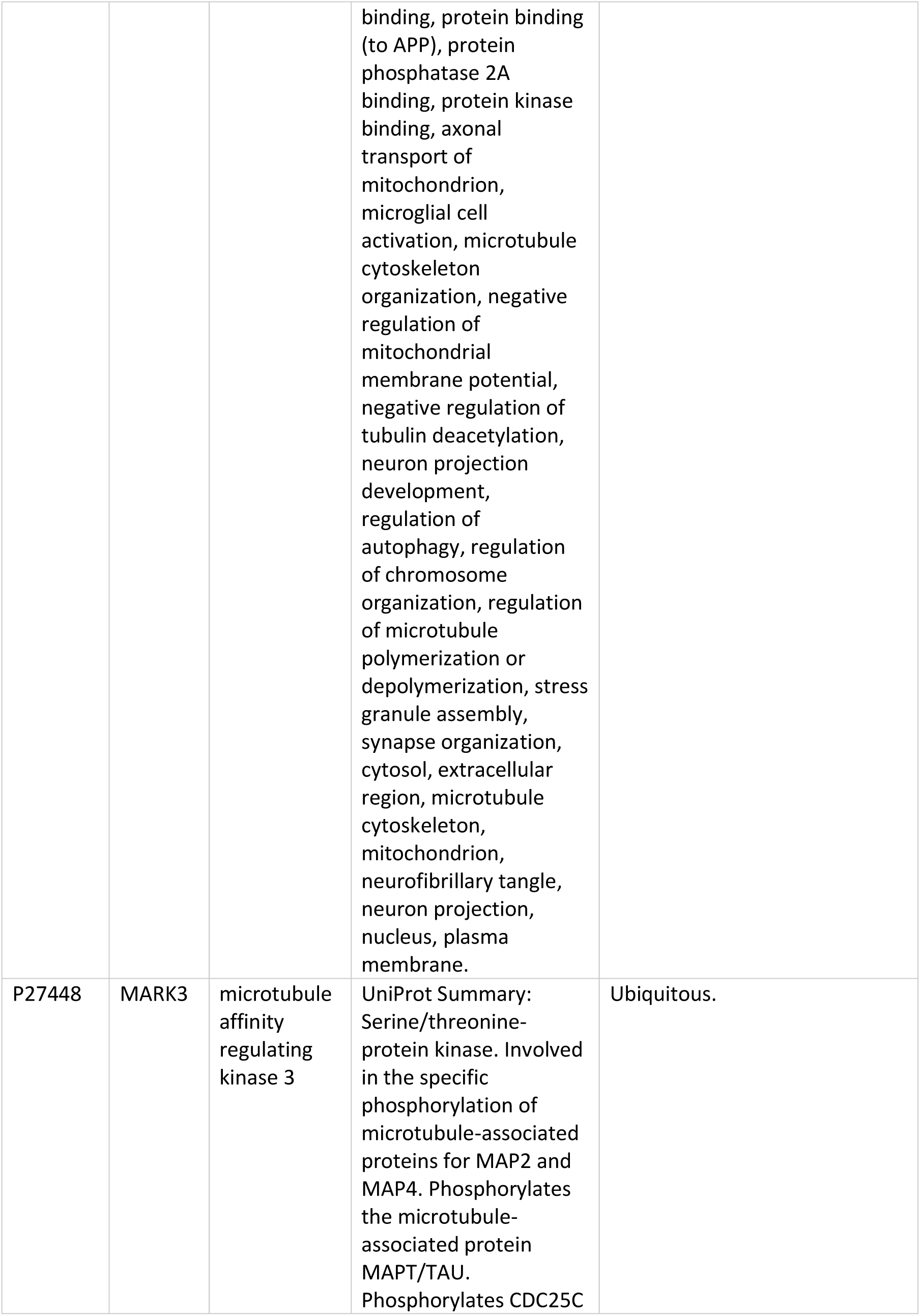
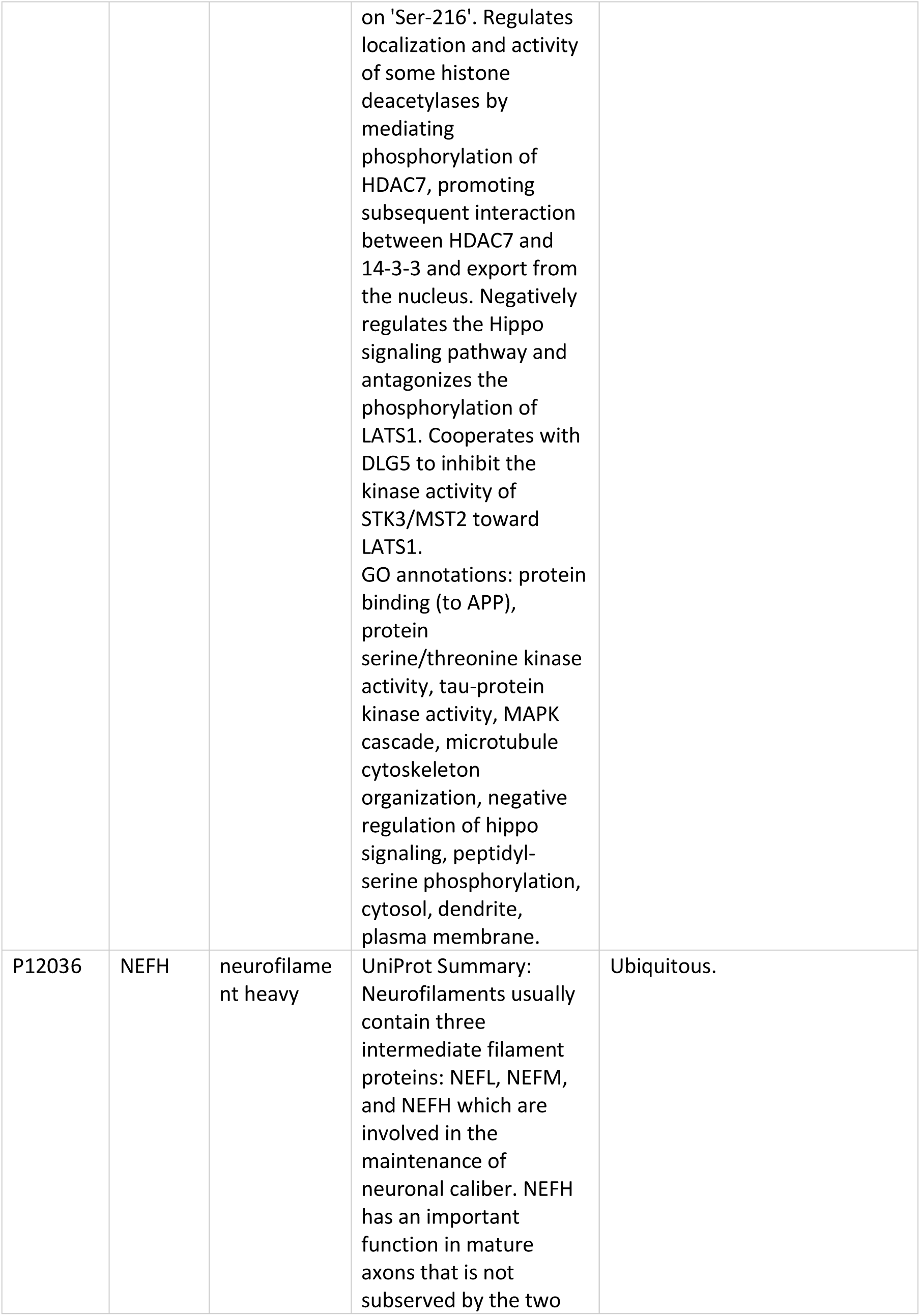
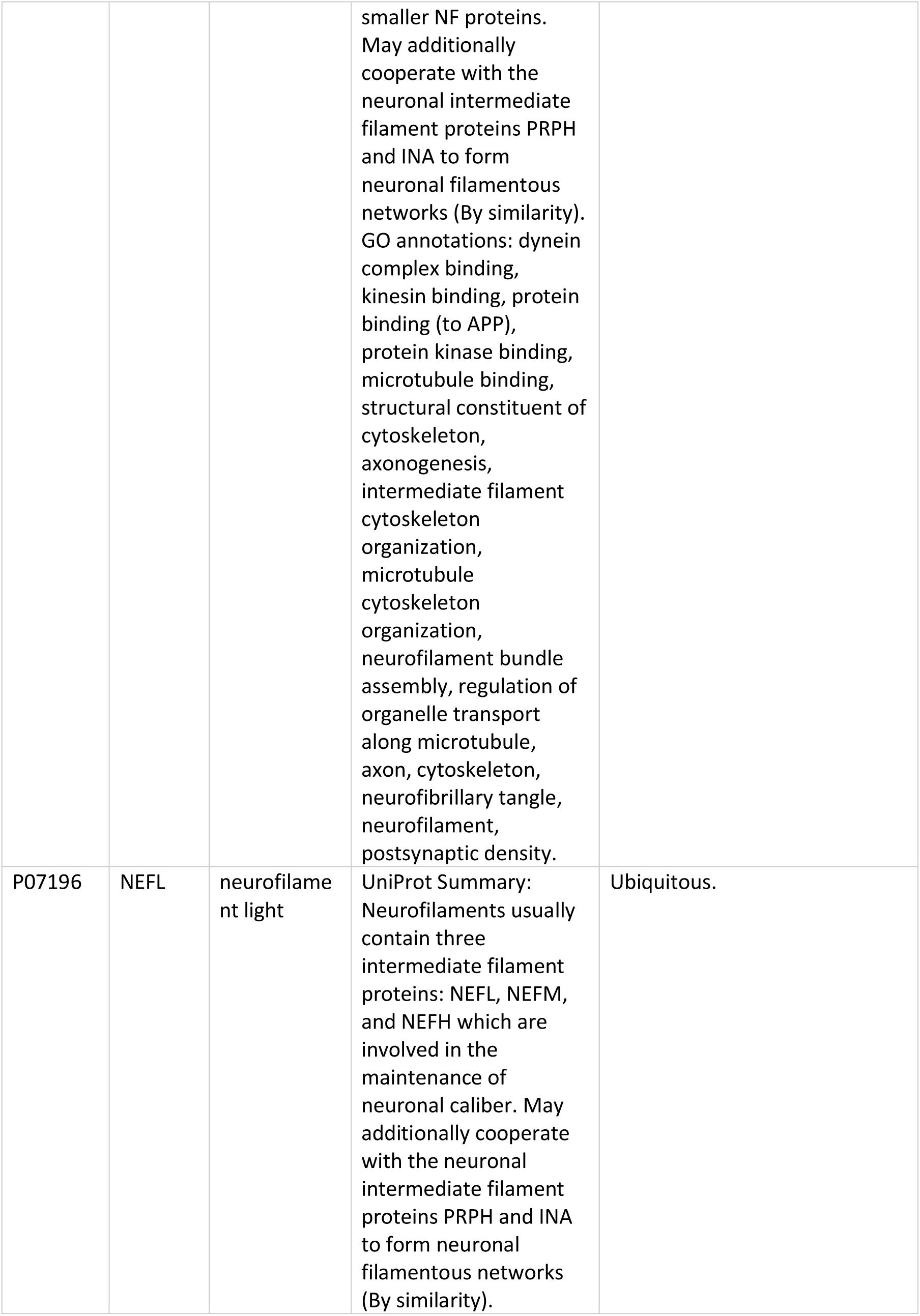
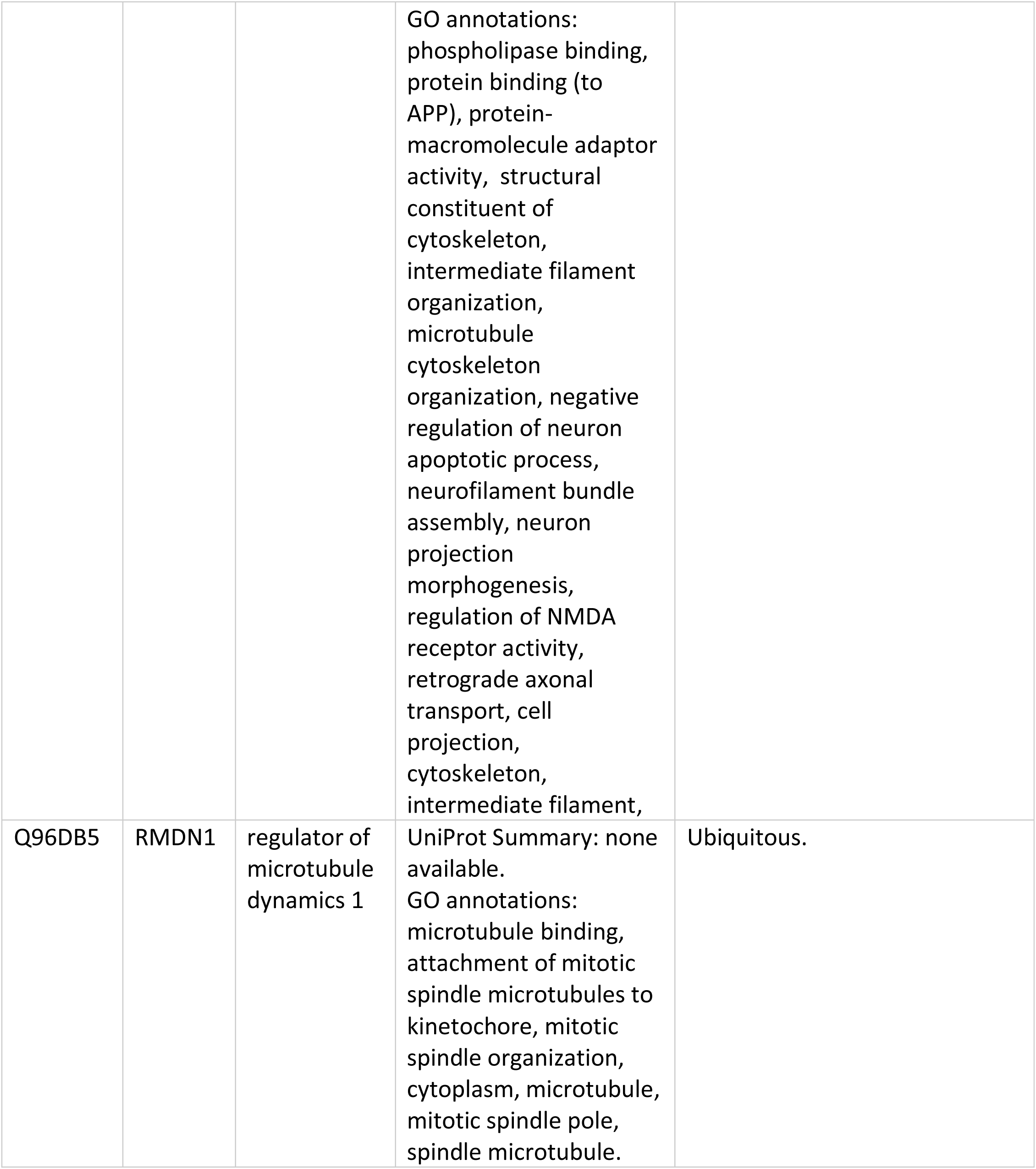
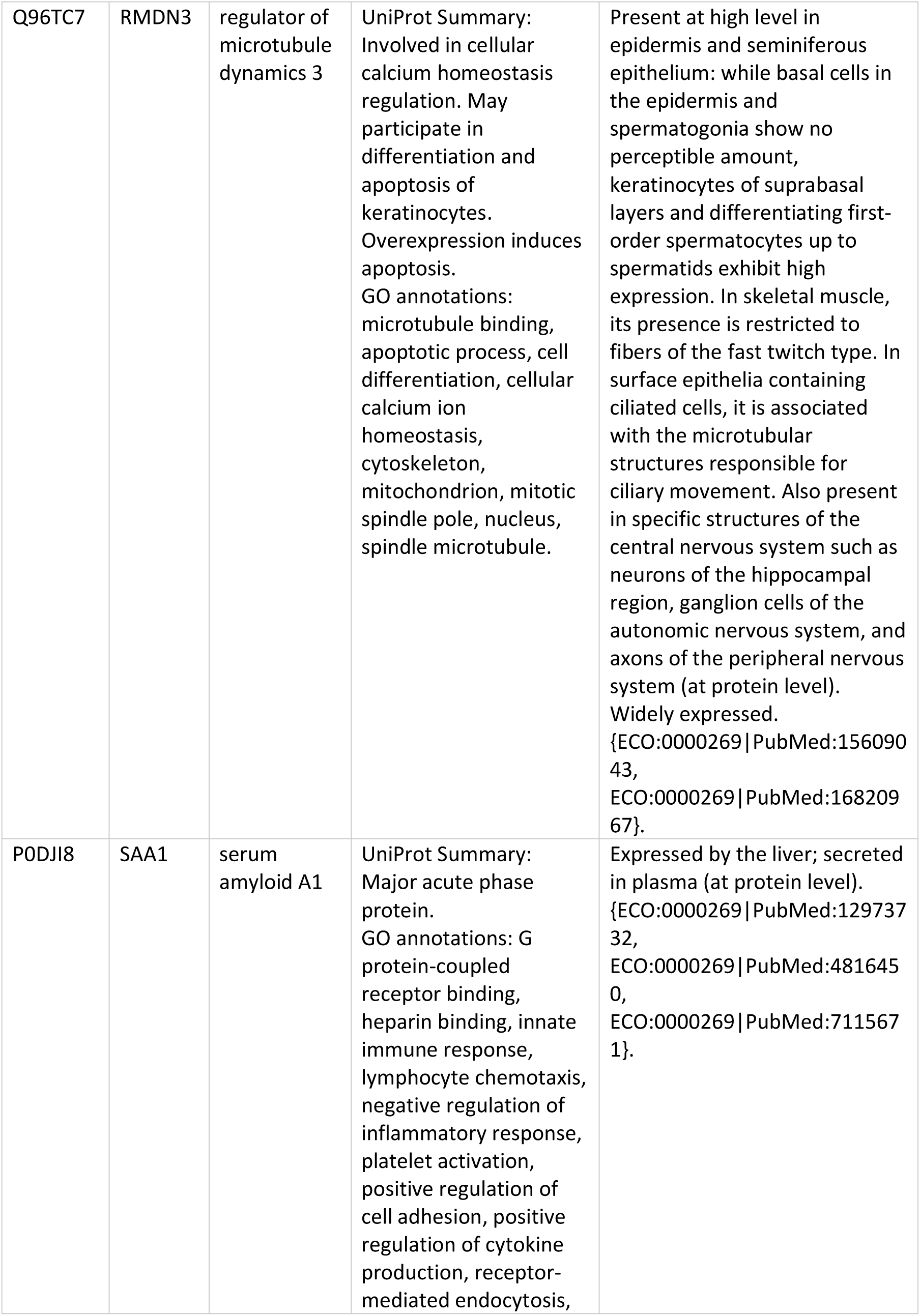
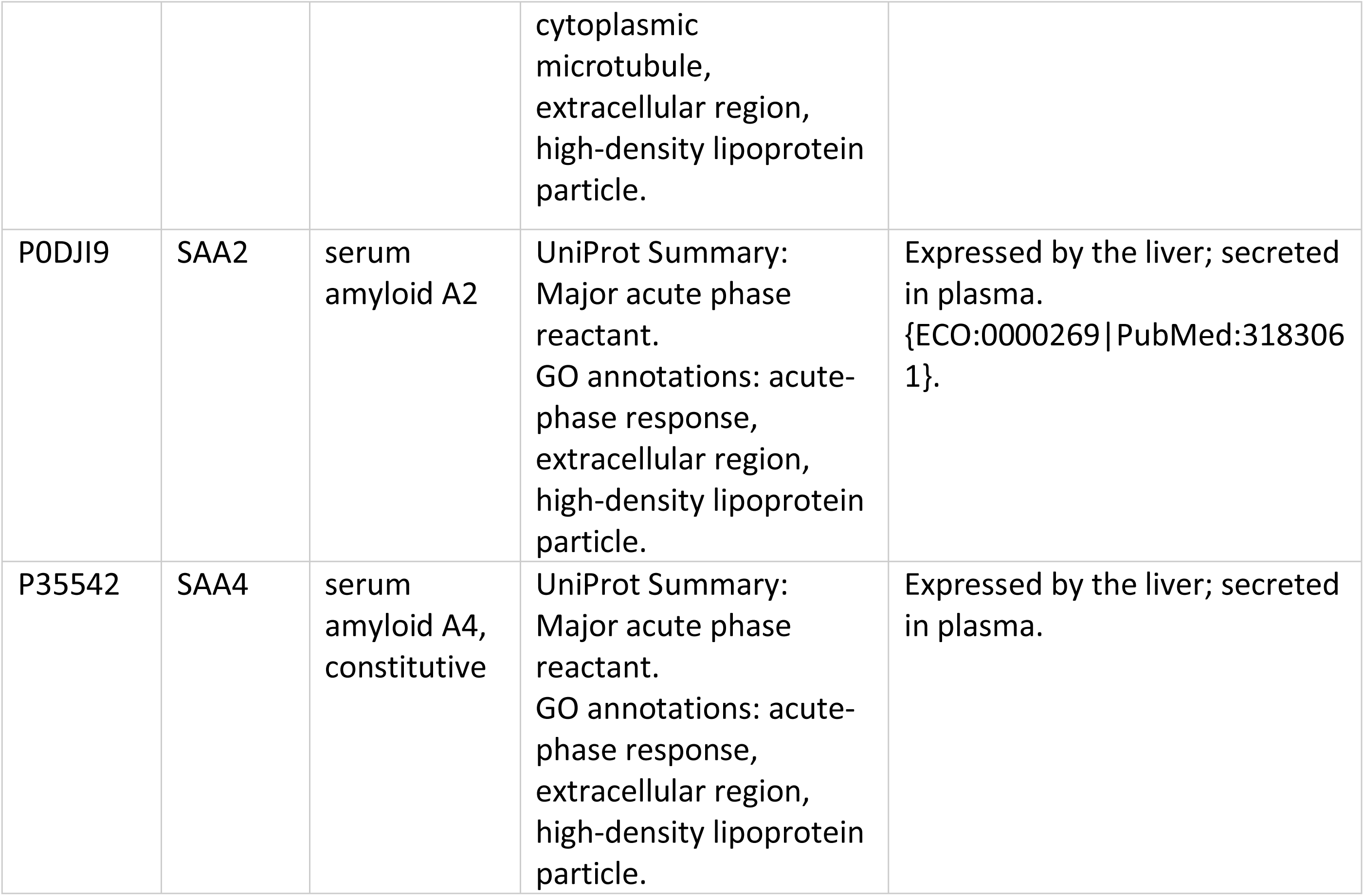
Amyloid, tau, and neurofilament related protein functions, tissue expression, and Gene Ontology terms from UniProt database.

**Supplementary Figure 1.**
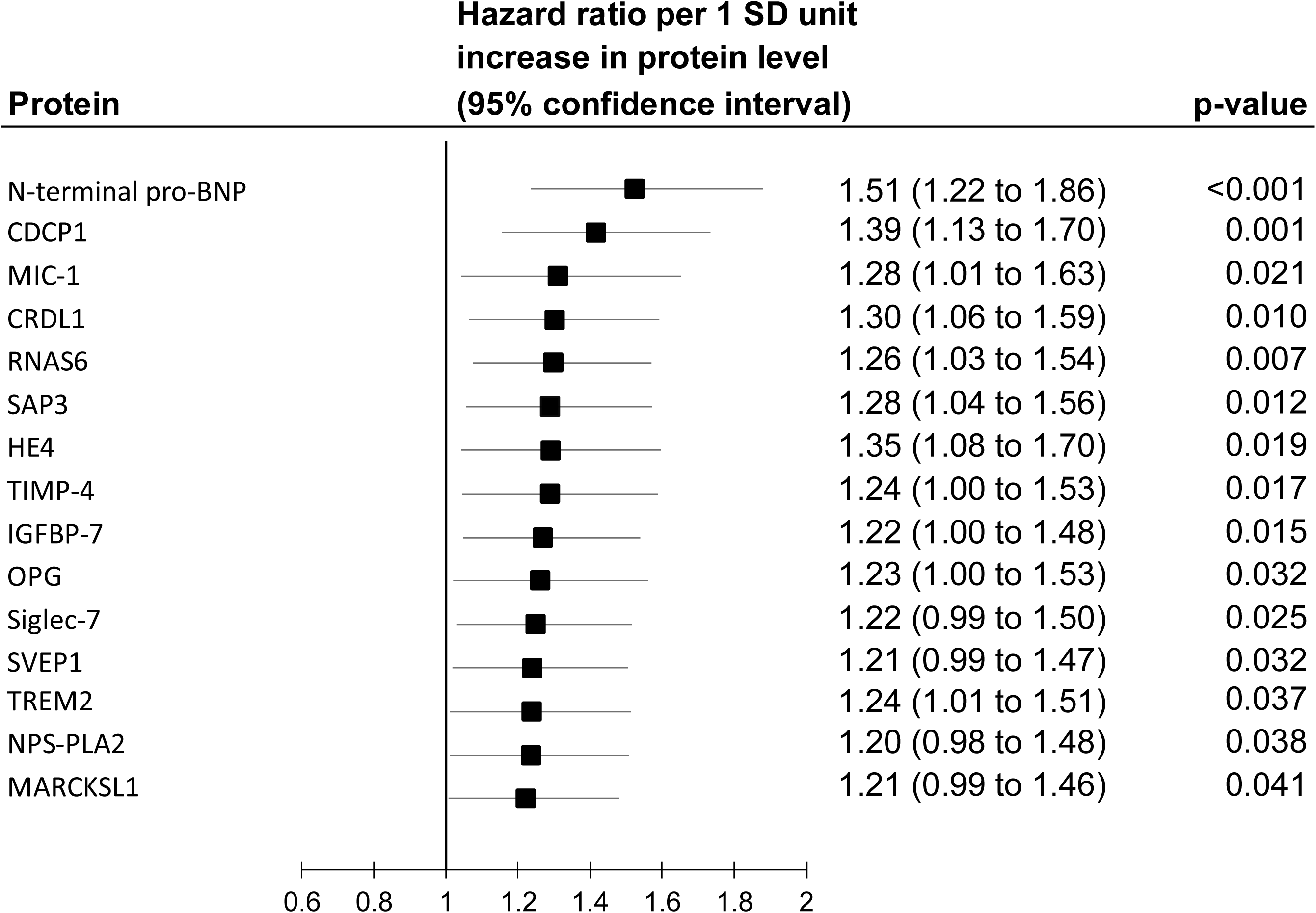
Age and sex adjusted results for the 15 proteins associated with both cognitive decline and dementia in the Whitehall II study.

**Supplementary Figure 2.**
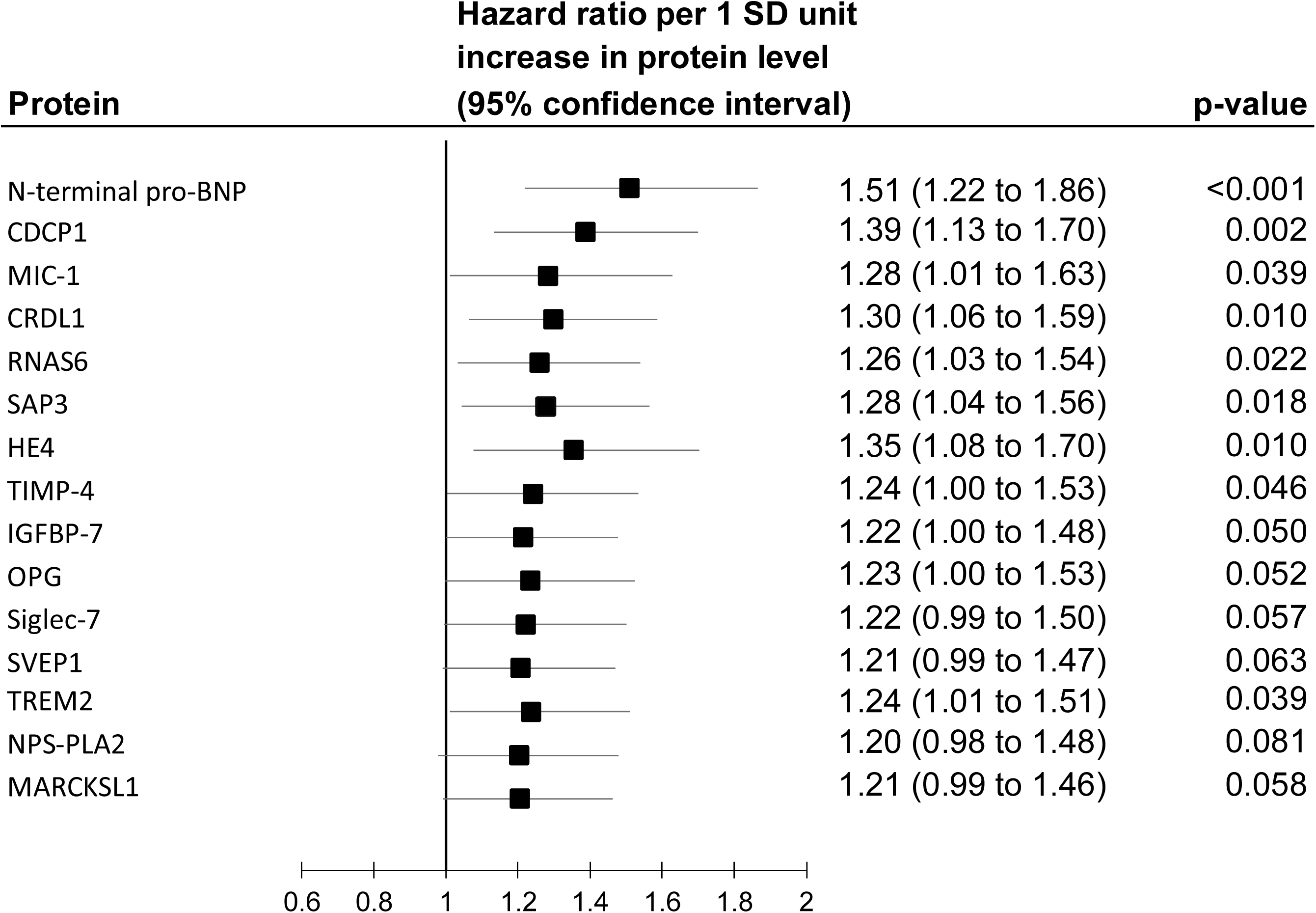
Age, sex, SBP, antihypertensive medication, smoking, diabetes, *APOE* genotype, BMI, alcohol consumption, and socioeconomic status adjusted results for the 15 proteins associated with both cognitive decline and dementia in the Whitehall II study.

**Supplementary Figure 3.**
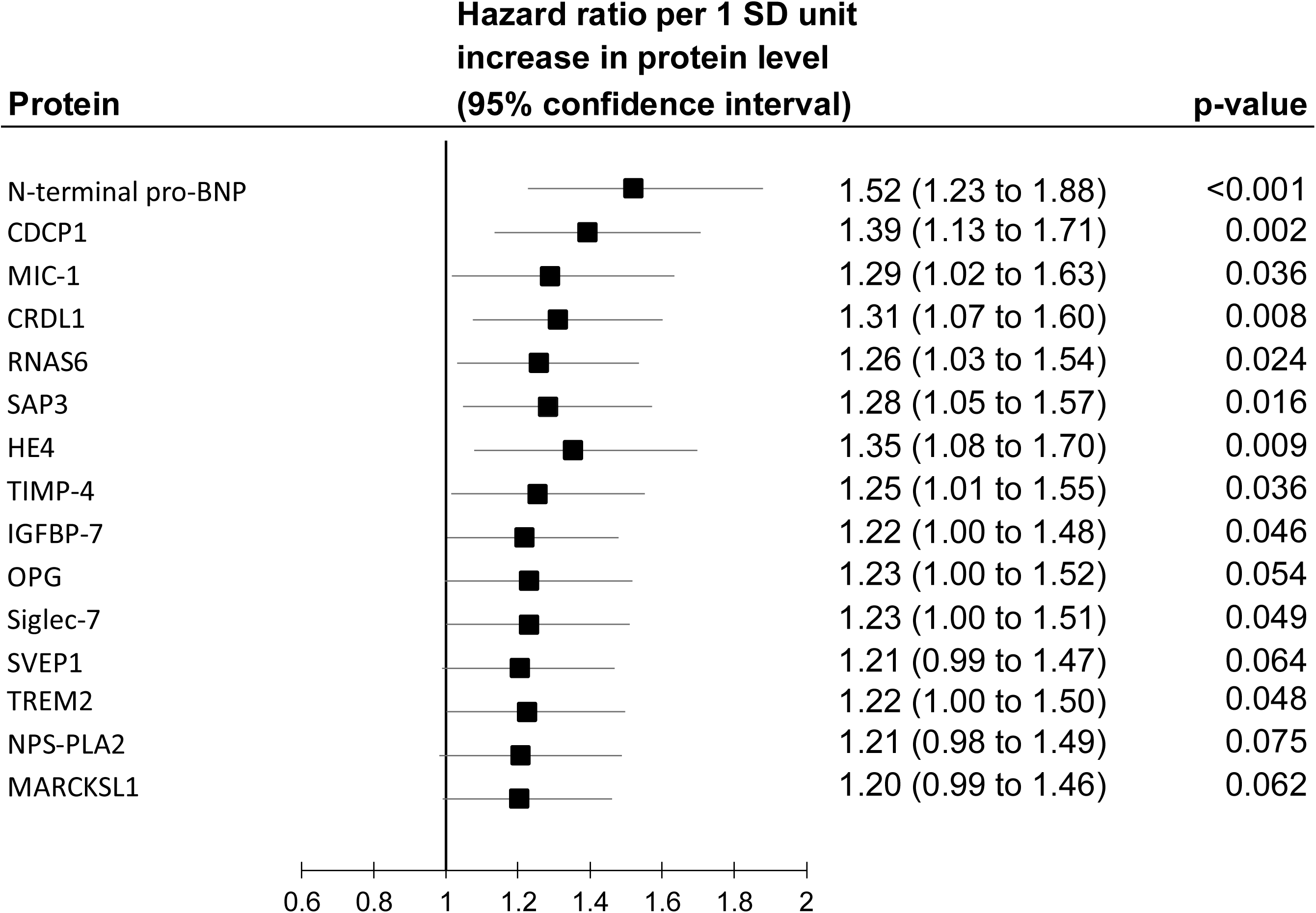
Age, sex, SBP, antihypertensive medication, smoking, diabetes, *APOE* genotype, BMI, alcohol consumption, and socioeconomic status adjusted results from competing risks (death) model for the 15 proteins associated with both cognitive decline and dementia in the Whitehall II study.

**Supplementary Table 3.** Associations between the cognitive decline slope and a SD increase in the 15 proteins that survived replication.

| **Protein** | **Beta (95% CIs)** | **p-value** | **FDR corrected p-value** |
| --- | --- | --- | --- |
| N-terminal pro-BNP | 0.12 (0.08 to 0.17) | 3E-09 | 2E-06 |
| CDCP1 | 0.16 (0.12 to 0.20) | 4E-14 | 1E-10 |
| MIC-1 | 0.18 (0.14 to 0.22) | 1E-17 | 7E-14 |
| CRDL1 | 0.09 (0.05 to 0.13) | 4E-05 | 0.0031 |
| RNAS6 | 0.07 (0.03 to 0.11) | 0.0007 | 0.023 |
| SAP3 | 0.08 (0.04 to 0.12) | 9E-05 | 0.0059 |
| HE4 | 0.12 (0.08 to 0.16) | 5E-09 | 2E-06 |
| TIMP-4 | 0.08 (0.04 to 0.13) | 7E-05 | 0.0050 |
| IGFBP-7 | 0.10 (0.06 to 0.14) | 2E-06 | 0.00040 |
| OPG | 0.12 (0.08 to 0.16) | 5E-09 | 2E-06 |
| SIGLEC-7 | 0.10 (0.05 to 0.14) | 6E-06 | 0.00084 |
| SVEP1 | 0.13 (0.09 to 0.17) | 2E-09 | 1E-06 |
| TREM2 | 0.09 (0.05 to 0.13) | 3E-05 | 0.0027 |
| NPS-PLA2 | 0.07 (0.03 to 0.11) | 0.0014 | 0.035 |
| MARCKSL1 | 0.07 (0.03 to 0.11) | 0.0006 | 0.021 |

**Supplementary Table 4.**
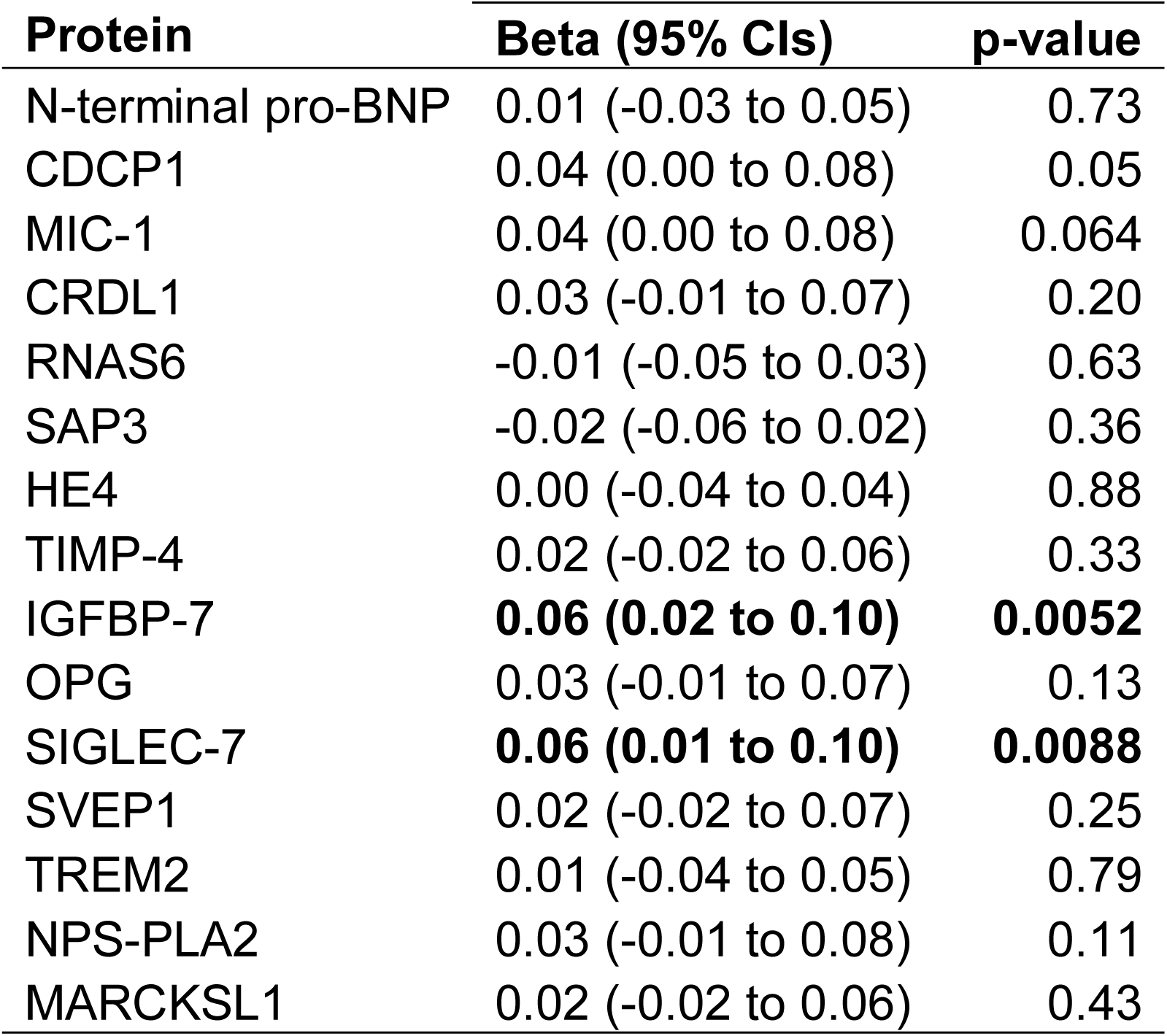
Associations between the memory decline slope and a SD increase in the 15 proteins that survived replication.

**Supplementary Table 5.** Associations between the phonemic fluency decline slope and a SD increase in the 15 proteins that survived replication.

| **Protein** | **Beta (95% CIs)** | **p-value** |
| --- | --- | --- |
| N-terminal pro-BNP | **0.10 (0.06 to 0.14)** | **1E-06** |
| CDCP1 | **0.08 (0.04 to 0.12)** | **8E-05** |
| MIC-1 | **0.08 (0.04 to 0.12)** | **0.0003** |
| CRDL1 | **0.04 (0.00 to 0.08)** | **0.046** |
| RNAS6 | 0.03 (-0.02 to 0.07) | 0.22 |
| SAP3 | 0.04 (0.00 to 0.08) | 0.058 |
| HE4 | **0.05 (0.01 to 0.09)** | **0.013** |
| TIMP-4 | **0.06 (0.02 to 0.10)** | **0.0053** |
| IGFBP-7 | **0.07 (0.02 to 0.11)** | **0.002** |
| OPG | **0.05 (0.01 to 0.09)** | **0.021** |
| SIGLEC-7 | 0.03 (-0.01 to 0.07) | 0.13 |
| SVEP1 | **0.09 (0.05 to 0.13)** | **2E-05** |
| TREM2 | **0.06 (0.02 to 0.11)** | **0.0024** |
| NPS-PLA2 | 0.02 (-0.02 to 0.06) | 0.33 |
| MARCKSL1 | **0.06 (0.02 to 0.10)** | **0.0069** |

**Supplementary Table 6.**
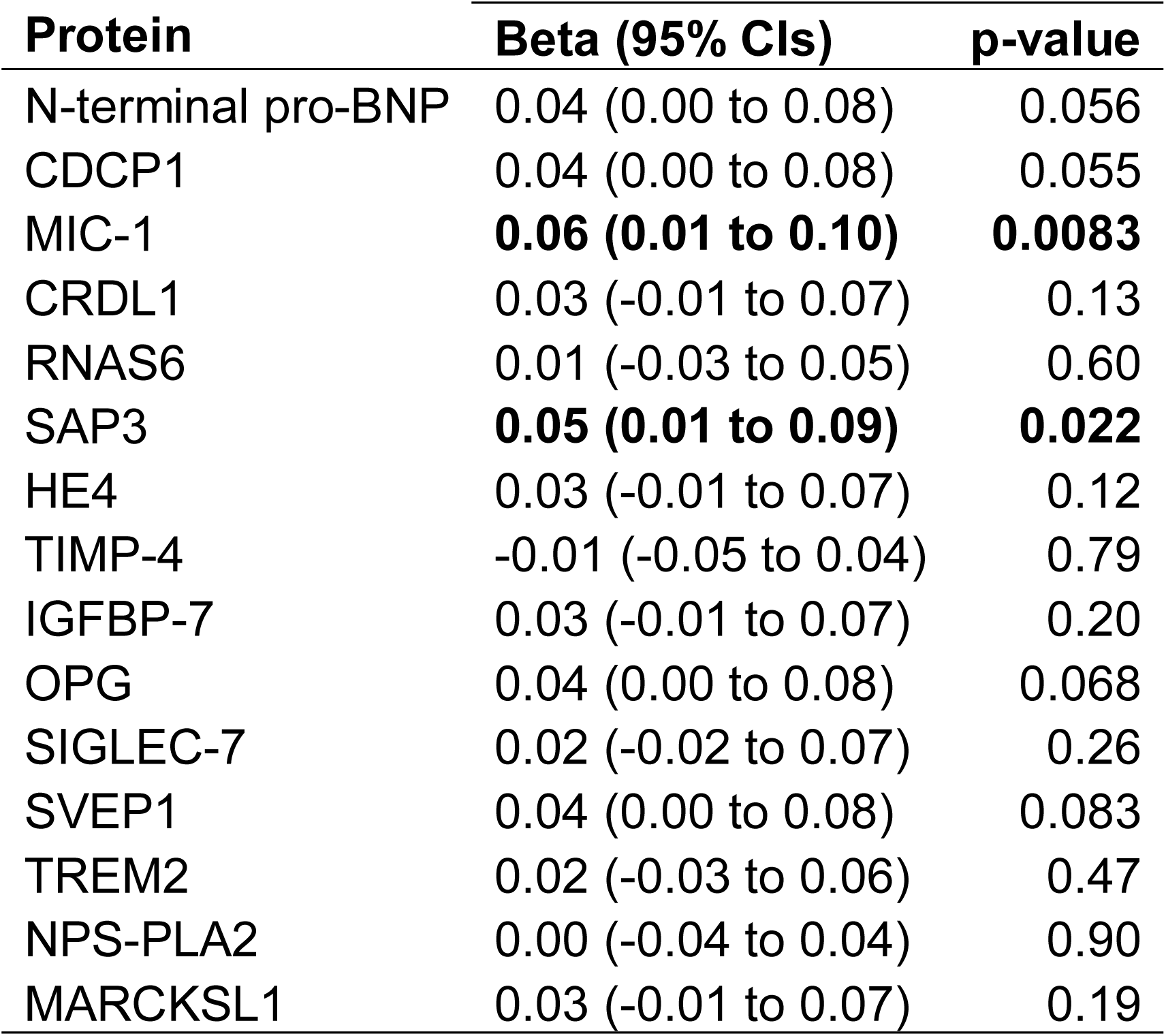
Associations between the semantic fluency decline slope and a SD increase in the 15 proteins that survived replication.

**Supplementary Table 7.** Associations between the executive function decline slope and SD increase in the 15 proteins that survived replication.

| **Protein** | **HR (95% CIs)** | **p-value** |
| --- | --- | --- |
| N-terminal pro-BNP | 0.14 (0.10 to 0.18) | 3E-11 |
| CDCP1 | 0.17 (0.12 to 0.21) | 3E-15 |
| MIC-1 | 0.20 (0.16 to 0.24) | 9E-21 |
| CRDL1 | 0.08 (0.04 to 0.12) | 8E-05 |
| RNAS6 | 0.10 (0.06 to 0.14) | 3E-06 |
| SAP3 | 0.10 (0.06 to 0.14) | 3E-06 |
| HE4 | 0.15 (0.10 to 0.19) | 5E-12 |
| TIMP-4 | 0.11 (0.07 to 0.15) | 3E-07 |
| IGFBP-7 | 0.06 (0.02 to 0.10) | 0.0054 |
| OPG | 0.14 (0.10 to 0.18) | 3E-11 |
| SIGLEC-7 | 0.07 (0.03 to 0.11) | 0.0007 |
| SVEP1 | 0.13 (0.09 to 0.17) | 1E-09 |
| TREM2 | 0.09 (0.05 to 0.13) | 9E-06 |
| NPS-PLA2 | 0.09 (0.05 to 0.13) | 3E-05 |
| MARCKSL1 | 0.09 (0.04 to 0.13) | 5E-05 |

**Supplementary Figure 4.**
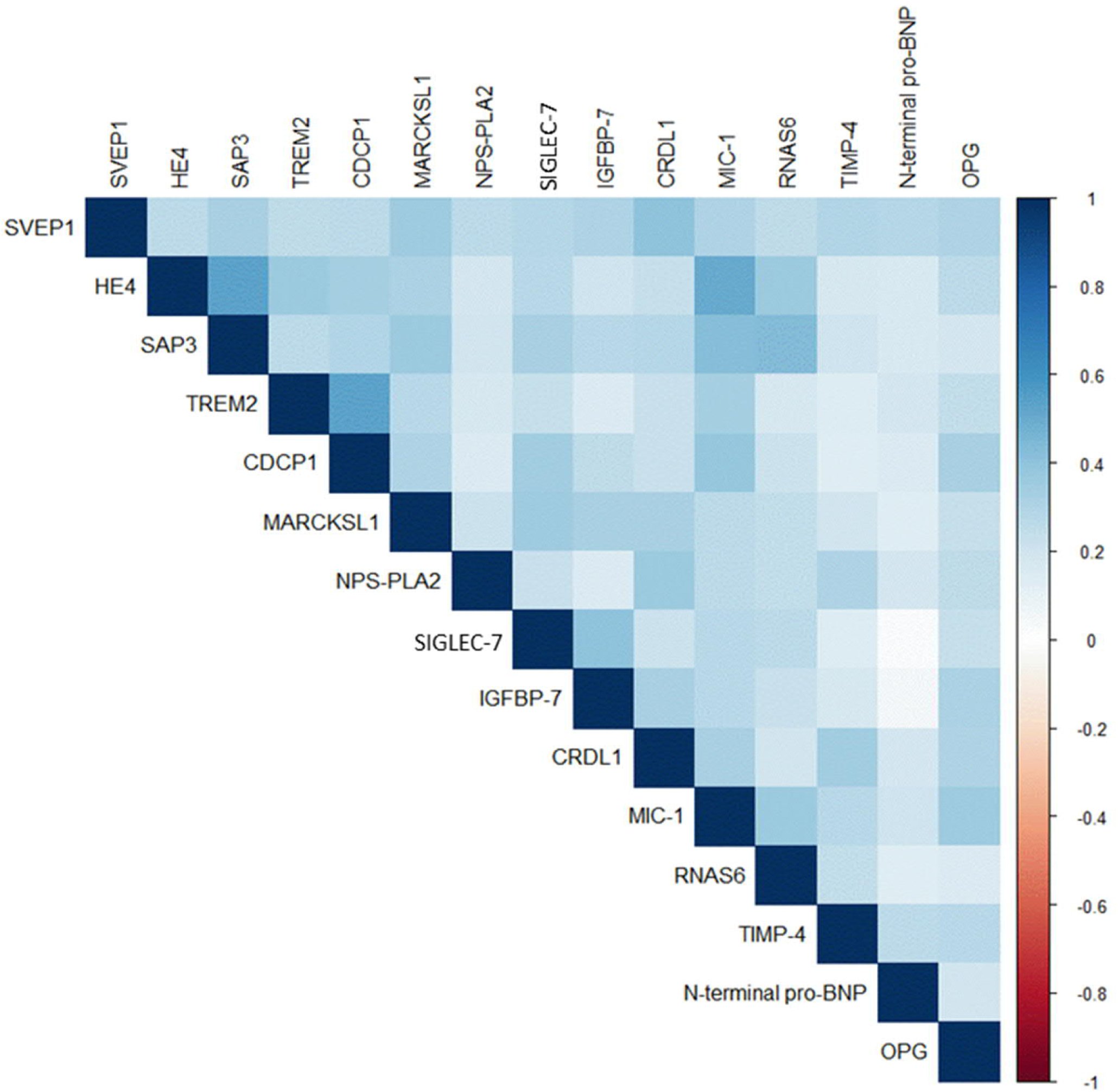
Between protein correlations.

**Supplementary Table 8.** Protein expression levels in the brain tissue for 15 proteins that survived replication.

| **Protein** | **Expression in brain** |
| --- | --- |
| N-terminal pro-BNP | Not known |
| CDCP1 | Low |
| MIC-1 | Not detected |
| CRDL1 | Not known |
| RNAS6 | Not detected |
| SAP3 | High |
| HE4 | Low |
| TIMP-4 | Not known |
| IGFBP-7 | Not known |
| OPG | Not known |
| SIGLEC-7 | Not known |
| SVEP1 | Low |
| TREM2 | Medium |
| NPS-PLA2 | Not detected |
| MARCKSL1 | Medium |

**Supplementary Table 9.** Drugs that can influence proteins associated with dementia, their potential mechanisms of action and current indications.

| Protein | Medication | Action | Indications |
| --- | --- | --- | --- |
| CDCP1 | Itolizumab | Prevents CDCP1 binding to CD6 and down regulates T cell activation and infiltration. It also reduces synthesis of pro-inflammatory cytokines reducing T cell infiltration at sites of inflammation. | Psoriasis |
| NPS-PLA2 | Varespladib and Varespladib Methyl | Inhibits arachidonic acid pathway in inflammation by inhibiting NPS-PLA2 activity and subsequent leukocyte activation. | Atherosclerotic cardiovascular diseases, inflammatory diseases, snake venom antidote |
| IGFBP-7 | Intranasal insulin, metformin and GLP-1 receptor agonists | Insulin nasal spray restores central insulin levels that may be down regulated by elevated IGFBP-7 levels. Metformin acts as insulin sensitizer and GLP-1 agonists stimulate insulin secretion. | Cognitive decline and Alzheimer's disease |
| N-terminal pro-BNP | Antihypertensive medications | Reduce N-terminal pro-BNP levels in circulation by reducing atrial and ventricular overload. | Hypertension |
| OPG | Atorvastatin, metformin, pioglitazone, rosiglitazone | Reduce OPG levels possibly by reducing inflammation and by stabilizing atherosclerotic plaques. | Atherosclerotic cardiovascular diseases, diabetes |

